## Supplementary material for "A modelling assessment of short- and medium-term risks of programme interruptions for *gambiense* human African trypanosomiasis in the DRC": S1 Text: Methods


This work is built on a series of studies on modelling and analyses of eliminating transmission of *gambiense* human African trypanosomiasis (gHAT) in the Democratic Republic of Congo (DRC). The series of studies includes a model fitting paper[1] and a model projections paper. To aid the reader of the present study, much of the same model information is provided here.

### S1.1 The compartmental gHAT model

The gHAT model we considered in this study is a variant "Model 4" of the Warwick model presented in the literature[1–4]. gHAT infections among hosts are described by Equation (1). Human hosts are modelled by the SEIIRS model with two infectious compartments, stage 1 disease, $I_{1H}$, and stage 2 disease, $I_{2H}$. Vectors are modelled by using compartments for appropriately modelling tsetse when used in a host-vector model with disease[3]. Pupal stage tsetse, $P_V$, emerge into unfed susceptible adults, $S_V$, and following a blood-meal become either exposed, $E_V$, or have reduced susceptibility to the *Trypanosoma brucei gambiense* parasites, $G_V$ - this effect is known as the teneral phenomenon. Following an infection, tsetse have an extrinsic incubation period (EIP) before becoming onwardly infectious. To incorporate a more realistic EIP distribution, there are three exposed classes, $E_{1V}, E_{2V}, E_{3V}$, which result in a gamma-distributed EIP (rather than an exponential with only one).

In order to reduce the dimensionality of our ODE system (by one), the vector equations are non-dimensionalised using the scaling $N_H/N_V$, where $N_H$ is the total human population, and $N_V$ is the tsetse population size. This results in a new non-dimensionalised parameter, $m_{\text{eff}}$, which is $\frac{p_H N_V}{N_H}$ appearing in host equations ($p_H$ is the probability of a human being infected by a single infectious bloodmeal) and is referred to as the *effective vector density*.

The proportion of tsetse bites taken on low-risk and high-risk humans are $f_1$ and $f_4$, depending on the relative availability/attractiveness and the relative abundance of two risk groups. High-risk humans are assumed to be $r$-fold more likely to receive bites, i.e. $s_1 = 1$ and $s_4 = r$. Therefore, $f_i$'s can be calculated using $f_i = \frac{s_i N_{Hi}}{\sum_j s_j N_{Hj}}$.

$$
\text{Humans}\quad\left\{
\begin{aligned}
\frac{\mathrm{d}S_{Hi}}{\mathrm{d}t} &= \mu_H N_{Hi} + \omega_H R_{Hi} - \alpha m_{\mathrm{eff}} f_i \frac{S_{Hi}}{N_{Hi}} I_V - \mu_H S_{Hi}\\
\frac{\mathrm{d}E_{Hi}}{\mathrm{d}t} &= \alpha m_{\mathrm{eff}} f_i \frac{S_{Hi}}{N_{Hi}} I_V - (\sigma_H + \mu_H) E_{Hi}\\
\frac{\mathrm{d}I_{1Hi}}{\mathrm{d}t} &= \sigma_H E_{Hi} - (\varphi_H + \eta_H(Y) + \mu_H) I_{1Hi}\\
\frac{\mathrm{d}I_{2Hi}}{\mathrm{d}t} &= \varphi_H I_{1Hi} - (\gamma_H(Y) + \mu_H) I_{2Hi}\\
\frac{\mathrm{d}R_{Hi}}{\mathrm{d}t} &= \eta_H(Y) I_{1Hi} + \gamma_H(Y) I_{2Hi} - (\omega_H + \mu_H) R_{Hi}
\end{aligned}
\right.
$$

$$
\text{Tsetse}\quad\left\{
\begin{aligned}
\frac{\mathrm{d}P_V}{\mathrm{d}t} &= B_V N_H - (\xi_V + \tfrac{P_V}{K}) P_V\\
\frac{\mathrm{d}S_V}{\mathrm{d}t} &= \xi_V \mathbb{P}(\text{survive pupal stage}) P_V - \alpha S_V - \mu_V S_V\\
\frac{\mathrm{d}E_{1V}}{\mathrm{d}t} &= \alpha\big(1 - f_T(t)\big) p_V \big(\textstyle\sum_i f_i \dfrac{(I_{1Hi} + I_{2Hi})}{N_{Hi}} + f_A \dfrac{I_A}{N_A}\big)(S_V + \varepsilon G_V)\\
&\qquad -(3\sigma_V + \mu_V + \alpha f_T(t)) E_{1V}\\
\frac{\mathrm{d}E_{2V}}{\mathrm{d}t} &= 3\sigma_V E_{1V} - (3\sigma_V + \mu_V + \alpha f_T(t)) E_{2V}\\
\frac{\mathrm{d}E_{3V}}{\mathrm{d}t} &= 3\sigma_V E_{2V} - (3\sigma_V + \mu_V + \alpha f_T(t)) E_{3V}\\
\frac{\mathrm{d}I_V}{\mathrm{d}t} &= 3\sigma_V E_{3V} - (\mu_V + \alpha f_T(t)) I_V\\
\frac{\mathrm{d}G_V}{\mathrm{d}t} &= \alpha(1 - f_T(t))\Big(1 - p_V\big(\textstyle\sum_i f_i \dfrac{(I_{1Hi} + I_{2Hi})}{N_{Hi}} + f_A \dfrac{I_A}{N_A}\big)\Big) S_V\\
&\qquad -\alpha\Big(f_T(t) + (1 - f_T(t)) p_V \varepsilon \big(\textstyle\sum_i f_i \dfrac{(I_{1Hi} + I_{2Hi})}{N_{Hi}} + f_A \dfrac{I_A}{N_A}\big)\Big) G_V\\
&\qquad\quad -\mu_V G_V
\end{aligned}
\right.
\tag{1}
$$

### S1.2 Model assumptions and parameter estimates from MCMC fitting

Table A provides the estimates of fixed parameters available in the literature used in the previous gHAT model[1–3]. The parameters fitted during the model fitting are defined in Table B. Posterior distributions for these parameters are estimated using MCMC methods[1] and are available in our graphical user interface at https://hatmepp.warwick.ac.uk/fitting/v2/, or can be downloaded in Open Science Framework at https://osf.io/ck3tr/.

**Table A. Model parametrisation (fixed parameters).** Notation, a brief description, and the used values for fixed parameters.

| Notation | Description | Value |
|---|---|---|
| $N_H$ | Total human population size in 2015 | Fixed for each health zone [5] |
| $\mu_H$ | Natural human mortality rate | $5.4795\times10^{-5}$ days$^{-1}$ [6] |
| $B_H$ | Total human birth rate | $=\mu_H N_H$ |
| $\sigma_H$ | Human incubation rate | 0.0833 days$^{-1}$ [7] |
| $\varphi_H$ | Stage 1 to 2 progression rate | 0.0019 days$^{-1}$ [8,9] |
| $\omega_H$ | Recovery rate or waning-immunity rate | 0.006 days$^{-1}$ [10] |
| Sens | Active screening diagnostic sensitivity | 0.91 [11] |
| $B_V$ | Tsetse birth rate | 0.0505 days$^{-1}$ [3] |
| $\xi_V$ | Pupal death rate | 0.037 days $^{-1}$ |
| $K$ | Pupal carrying capacity | $=111.09N_H$ [3] |
| $\mathbb{P}$(pupating) | Probability of pupating | 0.75 |
| $\mu_V$ | Tsetse mortality rate | 0.03 days$^{-1}$ [7] |
| $\sigma_V$ | Tsetse incubation rate | 0.034 days$^{-1}$ [12,13] |
| $\alpha$ | Tsetse bite rate | 0.333 days$^{-1}$ [14] |
| $p_V$ | Probability of tsetse infection per infectious bite | 0.065 [7] |
| $\varepsilon$ | Reduced non-teneral susceptibility factor | 0.05 [2] |
| $f_H$ | Proportion of blood-meals on humans | 0.09 [15] |
| disp$_{\text{act}}$ | Overdispersion parameter for active detection | $4\times10^{-4}$ [1] |
| disp$_{\text{pass}}$ | Overdispersion parameter for passive detection | $2.8\times10^{-5}$ [1] |

[1] Value of $B_V$ is chosen to maintain constant population size without interventions.
[2] Value of $K$ is chosen to reflect the observed bounce back rate.

**Table B. Model parametrisation (posteriors of fitted parameters).** Notation, a brief description, and representative percentiles of the posterior distributions for fitted parameters.

| Notation | Description | Posterior (median [95% CI]) | |
|---|---|---|---|
| | | **Kwamouth** | **Tandala** |
| $R_0$ | Basic reproduction number (NGM approach) | 1.09 [1.06, 1.14] | 1.009 [1.006, 1.014] |
| $r$ | Relative bites taken on high-risk humans | 6.61 [3.15, 10.75] | 2.04 [1.30, 4.26] |
| $k_1$ | Proportion of low-risk people | 0.90 [0.82, 0.95] | 0.95 [0.85, 0.99] |
| $\gamma_H^{\text{pre}}$ | Pre-1998 treatment rate from stage 2 (days$^{-1}$) | $1.72 \times 10^{-3}$ $[0.38, 4.88] \times 10^{-3}$ | $2.53 \times 10^{-3}$ $[1.03, 7.15] \times 10^{-3}$ |
| $\eta_H^{\text{post}}$ | Post-1998 treatment rate from stage 1 (days$^{-1}$) | $1.24 \times 10^{-4}$ $[0.60, 2.74] \times 10^{-4}$ | $2.74 \times 10^{-4}$ $[1.11, 4.99] \times 10^{-4}$ |
| $\gamma_H^{\text{post}}$ | Post-1998 treatment rate from stage 2 (days$^{-1}$) | $1.88 \times 10^{-3}$ $[0.46, 5.42] \times 10^{-3}$ | $3.60 \times 10^{-3}$ $[1.72, 8.98] \times 10^{-3}$ |
| Spec | Active screening diagnostic specificity | 0.9991 [0.9987, 0.9997] | 0.9998 [0.9997, 0.9999] |
| $u$ | Proportion of stage 2 passive cases reported | 0.27 [0.18, 0.40] | 0.39 [0.29, 0.51] |
| $d_{\text{change}}$ | Midpoint year for passive improvement | 2005.8 [2004.4, 2007.3] | – |
| $\eta_{H_{\text{amp}}}$ | Relative improvement in passive stage 1 detection rate | 2.52 [0.92, 5.46] | – |
| $\gamma_{H_{\text{amp}}}$ | Relative improvement in passive stage 2 detection rate | 0.51 [0.24, 0.97] | – |
| $d_{\text{steep}}$ | Speed of improvement in passive detection rate (years$^{-1}$) | 0.94 [0.68, 1.29] | – |

[1] See Equation (2) for improved passive detections formulated by $d_{\text{change}}$, $\eta_{H_{\text{amp}}}$, $\gamma_{H_{\text{amp}}}$ and $d_{\text{steep}}$.

### S1.3 Active screening

Screening data is aggregated by year and the exact dates and frequencies of conducting active screening (AS) are unknown, therefore some assumptions were made as to when AS takes place. Our model assumed only low-risk humans participate in AS and used the ratio of assumed number of people screened ($N_{AS}$) and the number of low-risk humans ($k_1 N_H$) to decide the frequency of AS each year. When screening numbers were smaller than potential participants ($N_{AS} < k_1 N_H$), a single AS event was assumed to take place at the beginning of those years. On the other hand, multiple AS events were evenly distributed over the time of the corresponding years, i.e. a second AS event in July when $k_1 N_H < N_{AS} \leq 2k_1 N_H$; a second AS event in May and a third AS event in September when $2k_1 N_H < N_{AS} \leq 3k_1 N_H$; etc.

### S1.4 Formulation and parametrisation of improved passive detections in Bandundu and Bas Congo

Previous analysis on provincial-level staged data (Lumbala *et al.* for 2000–2012[16] and WHO HAT Atlas data 2015–2016[17]) indicated that improved passive detection has happened across the former Bandundu province and in the former Bas Congo province[1]. Logistic functions shown in Equation (2) were used to formulate the improved passive detections in Bandundu and Bas Congo in year $Y$.

$$\begin{aligned}\eta_H(Y) &= \eta_H^{\text{post}} \left[ 1 + \frac{\eta_{H_{\text{amp}}}}{1 + \exp\left(-d_{\text{steep}}(Y - d_{\text{change}})\right)} \right], \\ \gamma_H(Y) &= \gamma_H^{\text{post}} \left[ 1 + \frac{\gamma_{H_{\text{amp}}}}{1 + \exp\left(-d_{\text{steep}}(Y - d_{\text{change}})\right)} \right].\end{aligned} \tag{2}$$

Parameter definitions and their posteriors are provided in Table B. N.B. It was assumed that improvements in both stages shared the same midpoint year and speed of improvement within a health zone. However, the amplitude of variation in each health zone came from the fitting of health-zone-specific data.

### S1.5 Formulation and parametrisation of additional tsetse mortality under vector control measures

The function which describes the probability of both hitting a target and dying is time dependent (days) from when the targets where placed:

$$f_T(t) = f_{\max} \left( 1 - \frac{1}{1 + \exp(-0.068(\text{mod}(t, 182.5) - 127.75))} \right), \tag{3}$$

and $f_{\max}$ is chosen such that the tsetse population after one year is at the observed/assumed percentage reduction. For the simplified model this is given by $f_{\max} = 0.0305$ for a 60% reduction, $f_{\max} = 0.0525$ for an 80% reduction, and $f_{\max} = 0.0750$ for a 90% reduction.

### S1.6 Simulations performed

Simulations were performed based on 1,000 model realisations. Observation uncertainty was considered by drawing ten random samples from the predicted mean dynamics for each set of parameters. A beta-binomial distribution in which an overdispersion parameter $\rho$ was introduced to the binomial distribution was used to account for larger variance than the binomial. The probability of obtaining $m$ successes out of $n$ trials with probability $p$ and overdispersion parameter $\rho$ is

$$\text{BetaBin}(m; n, p, \rho) = \frac{\Gamma(n+1)\Gamma(m+a)\Gamma(n-m+b)\Gamma(a+b)}{\Gamma(n-m+1)\Gamma(n+a+b)\Gamma(a)\Gamma(b)}, \tag{4}$$

where $a = p(1/\rho - 1)$ and $b = a(1-p)/p$.

Main observable outputs including active and passive cases each year were predicted by 10,000 samples. Unobservable outputs, such as new infections and the year of elimination of transmission, were predicted directly from the 1,000 model realisations without sampling (parameter uncertainty but no observation uncertainty). Our model also has the capability of outputting unreported deaths and person years spent in stage 1 and stage 2.

### S1.7 Uncertainty

Model predictions propagate parameter and observation uncertainty (see above). We tried to represent this uncertainty in a variety of ways:

- Time series box plots were used to display statistical summaries of model predictions – the median (the middle line in each box), the lower and upper quartiles (the edges of each box showing 50% prediction intervals) and 95% prediction intervals (extended whiskers containing the middle 95% of outputs).
- The median year of elimination of transmission was used to indicate the estimated elimination year for a series of model predictions because neither extreme values (outliers) nor truncation of simulation will affect the estimates.

### S1.8 Proxy for elimination of transmission

The gHAT model we used here is a deterministic model described by ODEs with transition rates between compartments. In the deterministic model, variables such as new infections, new cases and deaths can be non-integer and their values are continuous. The stochastic model, on the other hand, has dynamics driven by randomly occurring events with associated probabilities and its variables capture the discrete nature of the population. Despite good agreements on the mean dynamics in both models, even at very low prevalence, the dynamics at the endgame are different. Because of the continuous nature of deterministic dynamics, the number of infected people asymptotes to zero rather than reaching it unlike the stochastic model. In this paper, an artificial elimination of transmission (EoT) threshold i.e. one new infection per health zone per year was applied to new infections to determine whether EoT has been achieved or not. Other values of EoT thresholds, such as one new infection per 100,000 or per 1,000,000 people per year, can be found in the literature[18,19]. Large variation in EoT threshold highlights the difficulty in choosing a proper threshold reflecting the reality. More detailed comparison between stochastic and deterministic model variants will be needed in the future to ensure robustness of year of EoT estimates arising from such a proxy threshold.

### S1.9 Model Updates

Variations on this “Warwick gHAT model” have been previously published, starting with Rock et al.[2] and were updated the based on more recent and geographically broad human case data[1,20]. The key difference between the model utilised in the present study and the recent papers[1,20] is the way it can generate new projections which allow for reduction in AS coverage, and lower rates of PS from both stage 1 and stage 2 for 2020 and 2021. There are no fundamental differences in the model itself, only the simulation of these different future scenarios.
