## Supplementary material for "A modelling assessment of short- and medium-term risks of programme interruptions for *gambiense* human African trypanosomiasis in the DRC": S2 Text: Additional model outputs

### A modelling assessment of short- and medium-term risks of programme interruptions for *gambiense* human African trypanosomiasis in the DRC
### Supporting information – S2 Text: Additional model outputs

**Ching-I Huang[1,2,+,*], Ronald E Crump[1,2,3], Emily H Crowley[1,2], Andrew Hope[4], Paul R Bessell[5], Chansy Shampa[6], Erick Mwamba Miaka[6], and Kat S Rock[1,2,+]**

[1]Zeeman Institute for System Biology and Infectious Disease Epidemiology Research, The University of Warwick, Coventry, U.K
[2]Mathematics Institute, The University of Warwick, Coventry, U.K.
[3]The School of Life Sciences, The University of Warwick, Coventry, U.K.
[4]Liverpool School of Tropical Medicine (LSTM), Liverpool, U.K.
[5]Independent Consultant, Edinburgh, U.K.
[6]Programme National de Lutte contre la Trypanosomiase Humaine Africaine (PNLTHA), Kinshasa, D.R.C.
[*]
[+]these authors contributed equally to this work

#### Additional results

Fig A shows summary information for the predicted total cases, new infections, disability-adjusted life years (DALYs, a measure of disease burden) and year of elimination of transmission (YEoT) for three example health zones under different interruption scenarios. This is similar to the information provided in main text Figs 2 – 4, however, Fig A displays the total impact on outputs in each health zone between 2020 to 2030 compared the time series as displayed in the main text. This figure focuses on the overall impact of the interruption scenarios.

Figs B – E show time series of model outputs for health zones that have started vector control (VC) prior to the COVID-19 pandemic, Figs F – J show those that had plans for VC from 2020 or 2021, and Figs K – AJ show those that didn't have plans for VC, respectively. Model outputs include estimated active and passive cases, underlying new infections, disability-adjusted life years (DALYs, a measure of disease burden), and predicted probability of elimination of transmission (PEoT). Across health zones with existing VC or VC plans, the trends of model outputs are similar during and after the interruption period.

During the interruption period, the general features of model outputs are:

- Fewer active cases but more DALYs
- More passive cases when PS is fully operational (*No AS* scenario only)
- Fewer passive cases when there is reduced levels of PS (*No AS and reduced PS* and *No AS or VC and reduced PS* scenarios)
- Little impact on new infections and PEoT if VC as planned (*No AS* and *No AS and reduced PS* scenarios)
- More new infections and lower PEoT if VC suspended or postponed (*No AS or VC and reduced PS* scenario)

After the interruption period, the general features of model outputs are

- More active and passive cases and DALYs for a few years compared to the baseline
- New infections decline rapidly in 1–2 years in the *No AS or VC and reduced PS* scenario in health zones with planned VC
- Achieving EoT (i.e. PEoT = 1) with a maximum delay of the length of interruption if it hasn't reach EoT prior to the COVID-19 pandemic

The Programme National de Lutte contre la Trypanosomiase Humaine Africaine (PNLTHA) in the DRC were not able to operate AS through their mobile screening teams for some months in 2020. The provincial-level VC teams managed to continue the scheduled Tiny Target deployment in five health zones with existing VC (i.e. Bandundu, Kikongo, Kwamouth, Masi Manimba, and Yasa Bonga) in 2020 and 2021[1] so the actual situation might be between the *No AS* and *No AS and reduced PS* scenarios. But all the planned VC in Bokoro, Bolobo, Bulungu, Kokala, and Mushie were postponed, which means the *No AS or VC, and reduced PS* scenario might be considered closer to the reality in these regions.

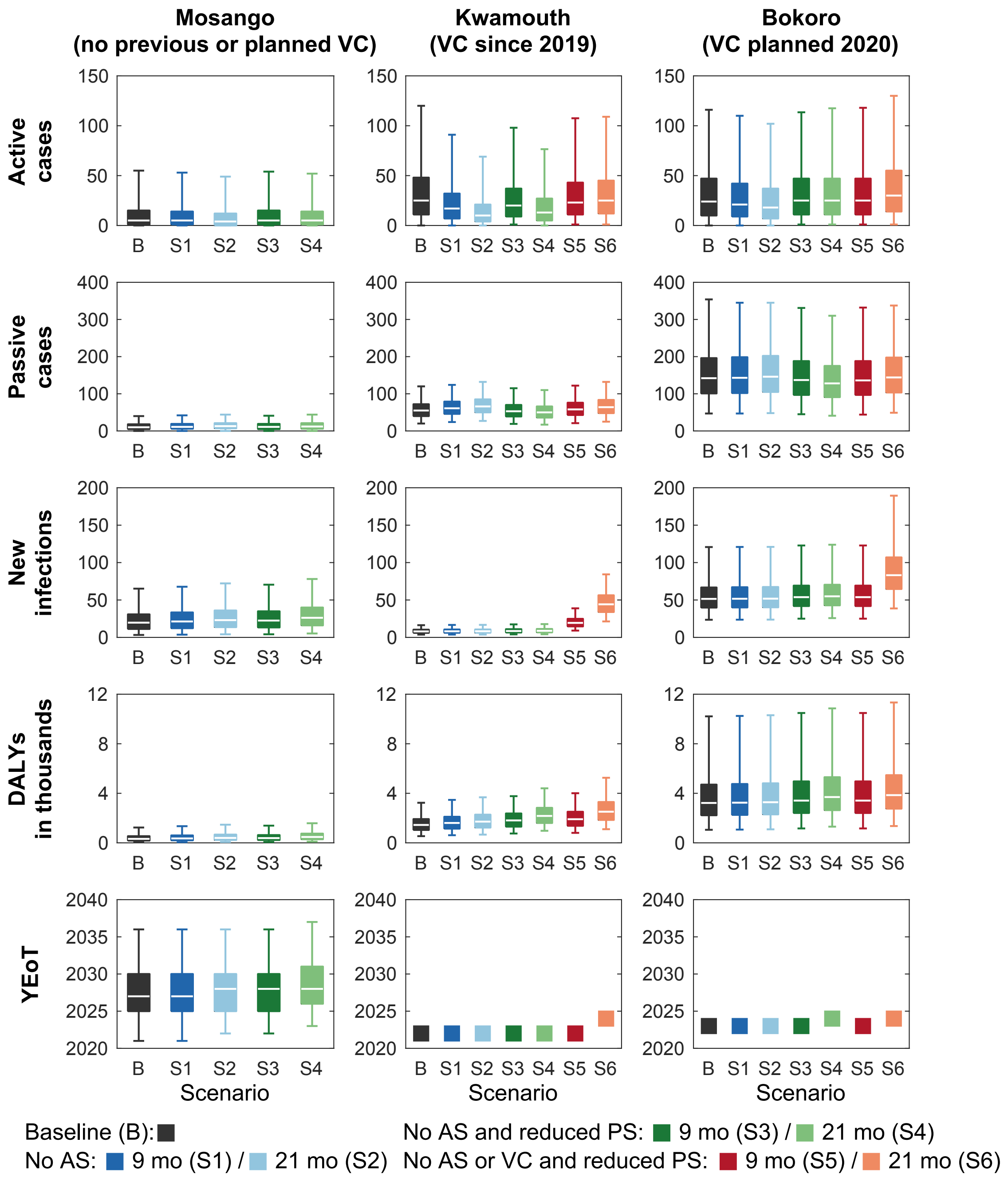


**Figure A. Model outputs for 2020–2030 in Mosango, Kwamouth and Bokoro health zones under the baseline and six interruption scenarios.** In Mosango only four interruption scenarios are shown as no VC is on-going or planned, therefore S5 and S6 are identical to S3 and S4, respectively. Box and whiskers show medians (centre line), 50% prediction intervals (PIs, box), and 95% PIs (whiskers). Black colours show the baseline (B) scenario, blues show *No AS* scenarios, greens show *No AS and reduced PS* scenarios and reds show *No AS or VC and reduced PS* scenarios. Darker colours show 9-month interruptions and lighter show 21-month interruptions. In the case of the expected year of elimination of transmission (YEoT) in Kwamouth and Bokoro the PIs are present but are very small therefore difficult to visualise.
AS: active screening; PS: passive screening; VC: vector control, DALYs: disability-adjusted life years, YEoT: year of elimination of transmission

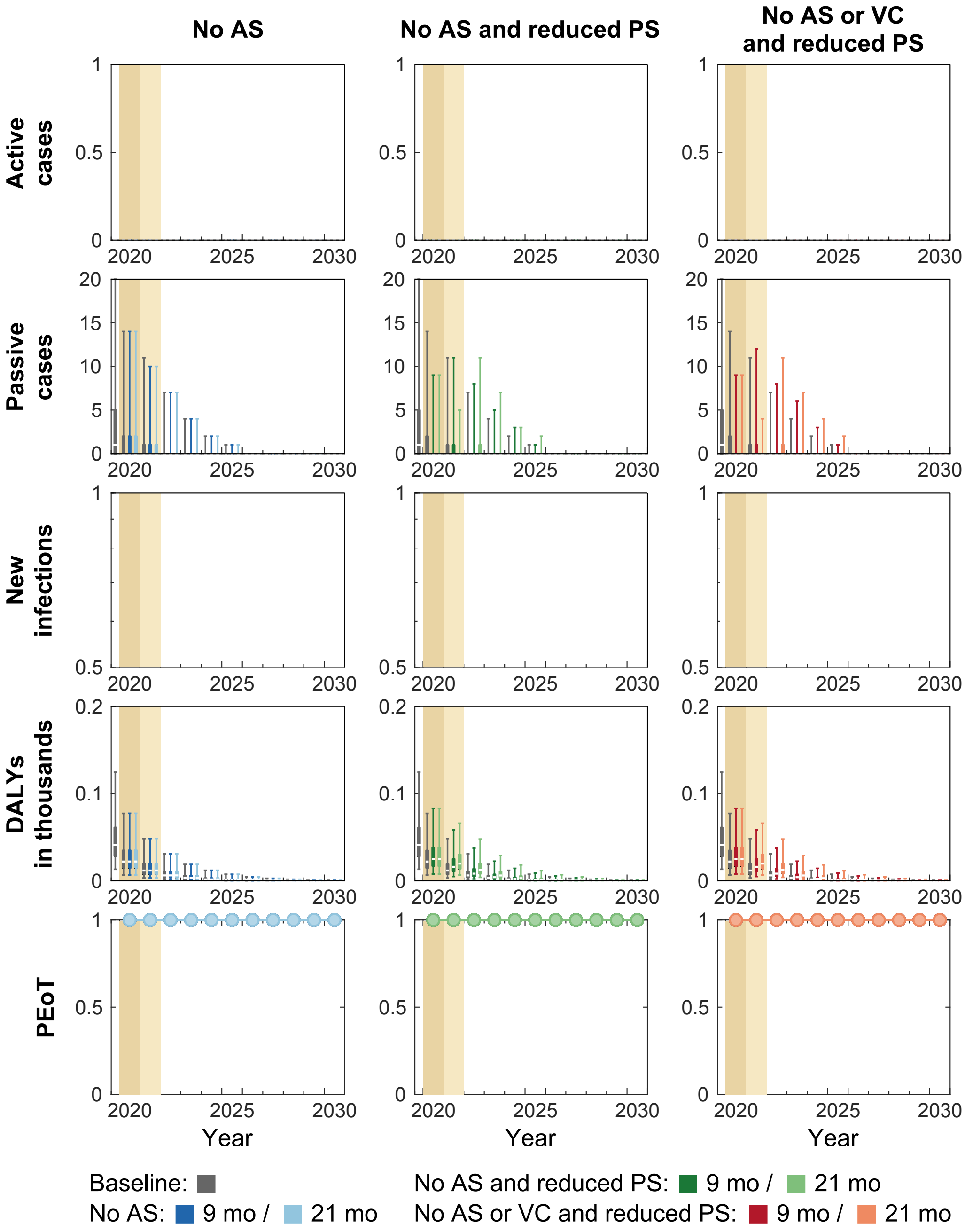


**Figure B. Time series of model outputs in Yasa Bonga health zone under the baseline and six interruption scenarios.** During 2014–2018, an average of 57% of the population participated in active screening resulting in 2.23 reported cases (from both active and passive screenings) per 10,000 per annum in Yasa Bonga health zone. The first Tiny Target deployment of vector control was in mid-2015. Interruptions by COVID-19 are assumed to take place in April 2020 and last until the end of 2020 or 2021 in our simulations. There are n = 10,000 independent samples, 10 from each of 1,000 independent samples from the joint posterior distributions of the fitted model parameters. Box plots summarise parameter and observational uncertainty. The lines in the boxes represent the medians of predicted results. The lower and upper bounds of the boxes indicate 25th and 75th percentiles. The minimum and maximum values are 2.5th and 97.5th percentiles and therefore whiskers cover 95% prediction intervals.

AS: active screening; PS: passive screening; VC: vector control; DALYs: disability-adjusted life years; PEoT: probability of elimination of transmission

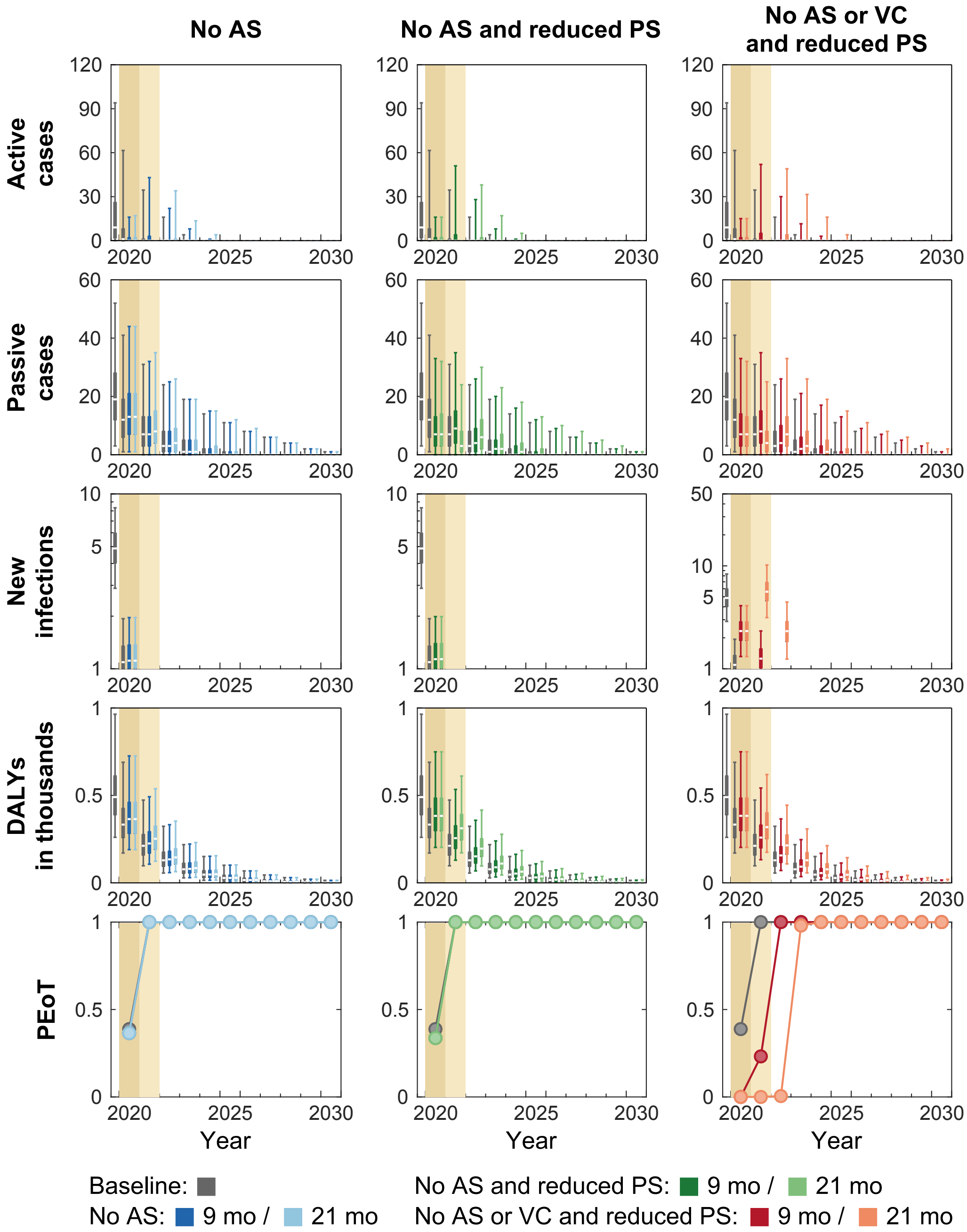


**Figure C. Time series of model outputs in Masi Manimba health zone under the baseline and six interruption scenarios.** During 2014–2018, an average of 46% of the population participated in active screening resulting in 2.98 reported cases (from both active and passive screenings) per 10,000 per annum in Masi Manimba health zone. The first Tiny Target deployment of vector control was in mid-2018. Interruptions by COVID-19 are assumed to take place in April 2020 and last until the end of 2020 or 2021 in our simulations. There are n = 10,000 independent samples, 10 from each of 1,000 independent samples from the joint posterior distributions of the fitted model parameters. Box plots summarise parameter and observational uncertainty. The lines in the boxes represent the medians of predicted results. The lower and upper bounds of the boxes indicate 25th and 75th percentiles. The minimum and maximum values are 2.5th and 97.5th percentiles and therefore whiskers cover 95% prediction intervals.

AS: active screening; PS: passive screening; VC: vector control; DALYs: disability-adjusted life years; PEoT: probability of elimination of transmission

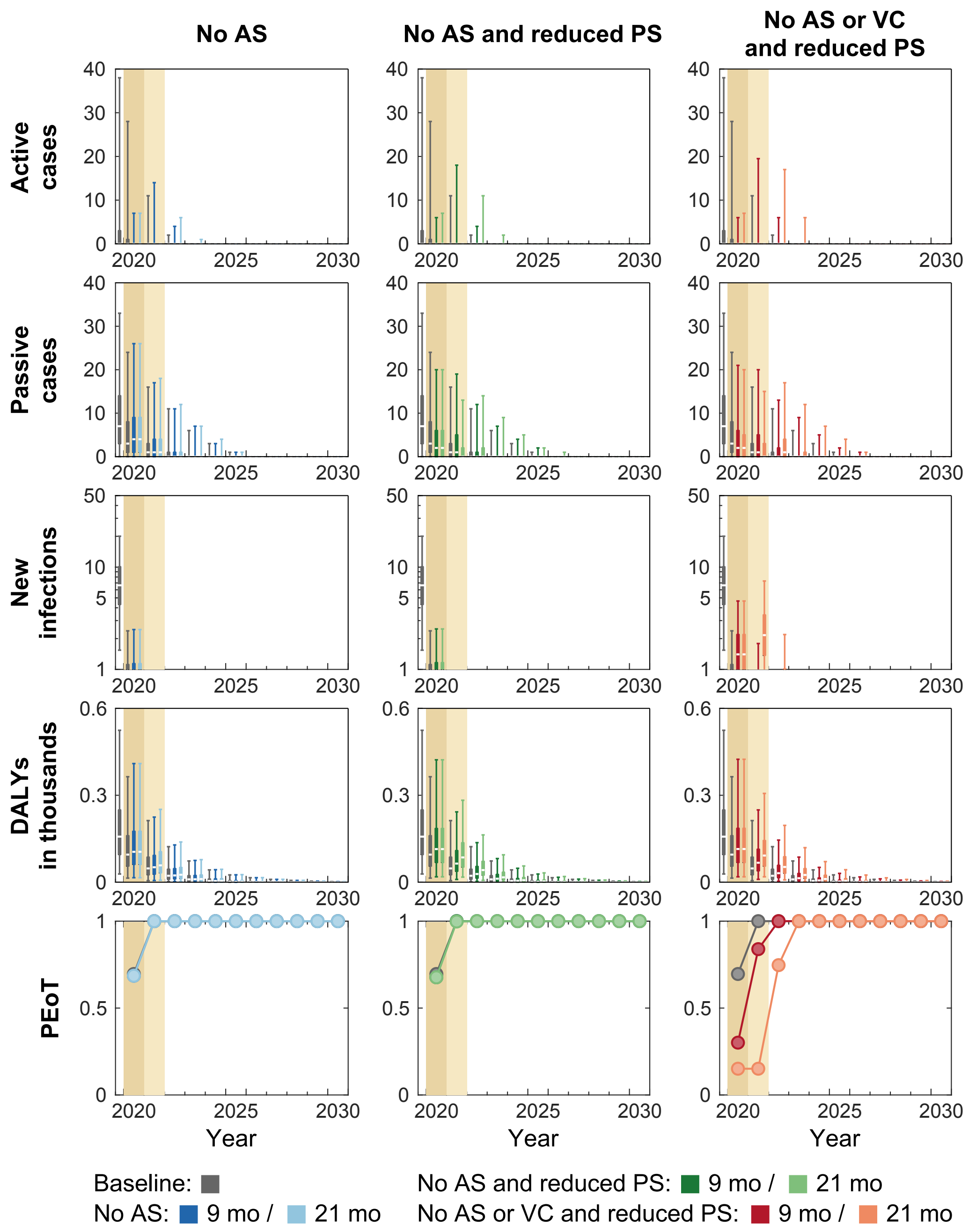


**Figure D. Time series of model outputs in Bandundu health zone under the baseline and six interruption scenarios.** During 2014–2018, an average of 34% of the population participated in active screening resulting in 2.75 reported cases (from both active and passive screenings) per 10,000 per annum in Bandundu health zone. The first Tiny Target deployment of vector control was in mid-2019. Interruptions by COVID-19 are assumed to take place in April 2020 and last until the end of 2020 or 2021 in our simulations. There are n = 10,000 independent samples, 10 from each of 1,000 independent samples from the joint posterior distributions of the fitted model parameters. Box plots summarise parameter and observational uncertainty. The lines in the boxes represent the medians of predicted results. The lower and upper bounds of the boxes indicate 25th and 75th percentiles. The minimum and maximum values are 2.5th and 97.5th percentiles and therefore whiskers cover 95% prediction intervals.

AS: active screening; PS: passive screening; VC: vector control; DALYs: disability-adjusted life years; PEoT: probability of elimination of transmission

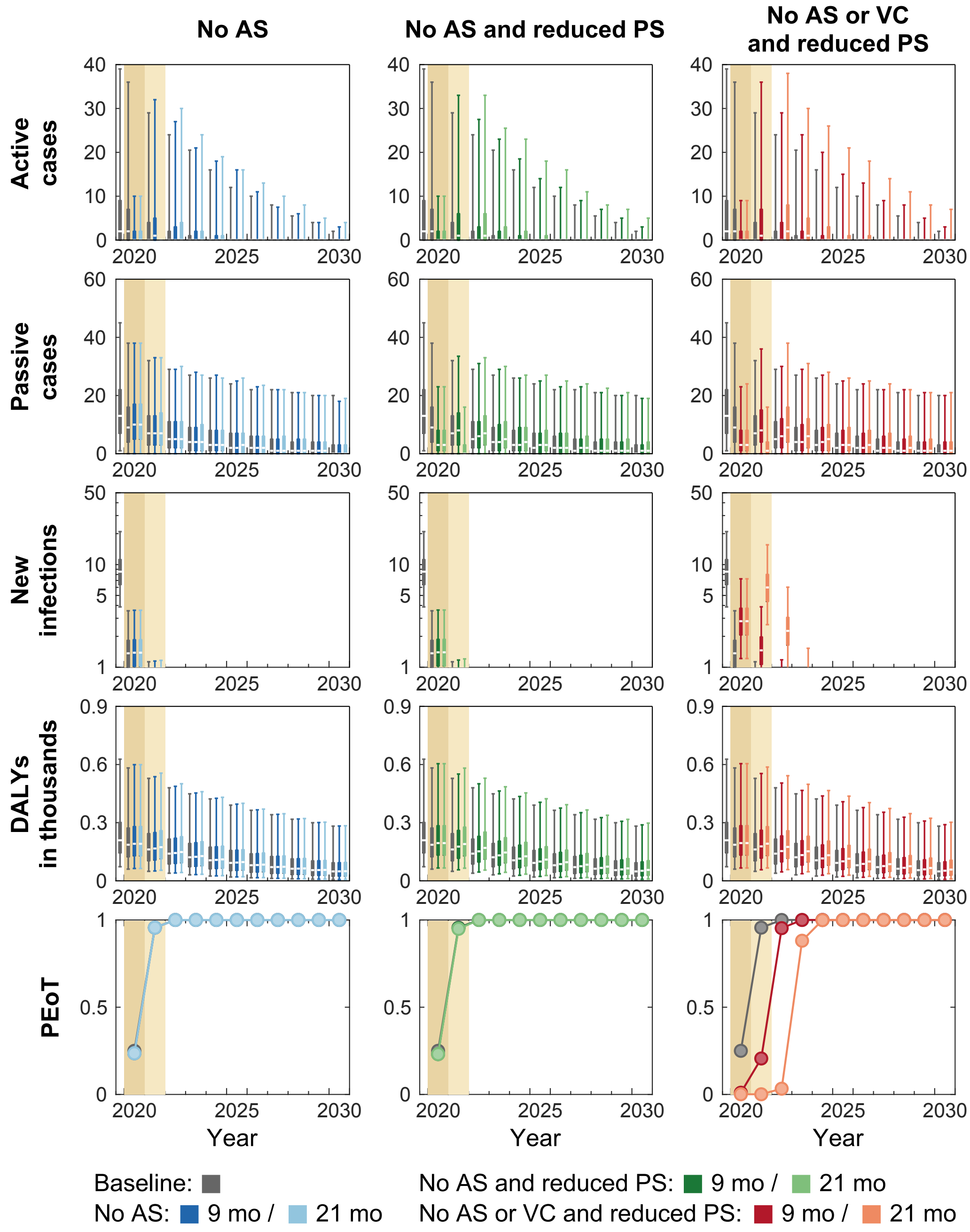


**Figure E. Time series of model outputs in Kikongo health zone under the baseline and six interruption scenarios.** During 2014–2018, an average of 21% of the population participated in active screening resulting in 2.82 reported cases (from both active and passive screenings) per 10,000 per annum in Kikongo health zone. The first Tiny Target deployment of vector control was in mid-2019. Interruptions by COVID-19 are assumed to take place in April 2020 and last until the end of 2020 or 2021 in our simulations. There are n = 10,000 independent samples, 10 from each of 1,000 independent samples from the joint posterior distributions of the fitted model parameters. Box plots summarise parameter and observational uncertainty. The lines in the boxes represent the medians of predicted results. The lower and upper bounds of the boxes indicate 25th and 75th percentiles. The minimum and maximum values are 2.5th and 97.5th percentiles and therefore whiskers cover 95% prediction intervals.

AS: active screening; PS: passive screening; VC: vector control; DALYs: disability-adjusted life years; PEoT: probability of elimination of transmission

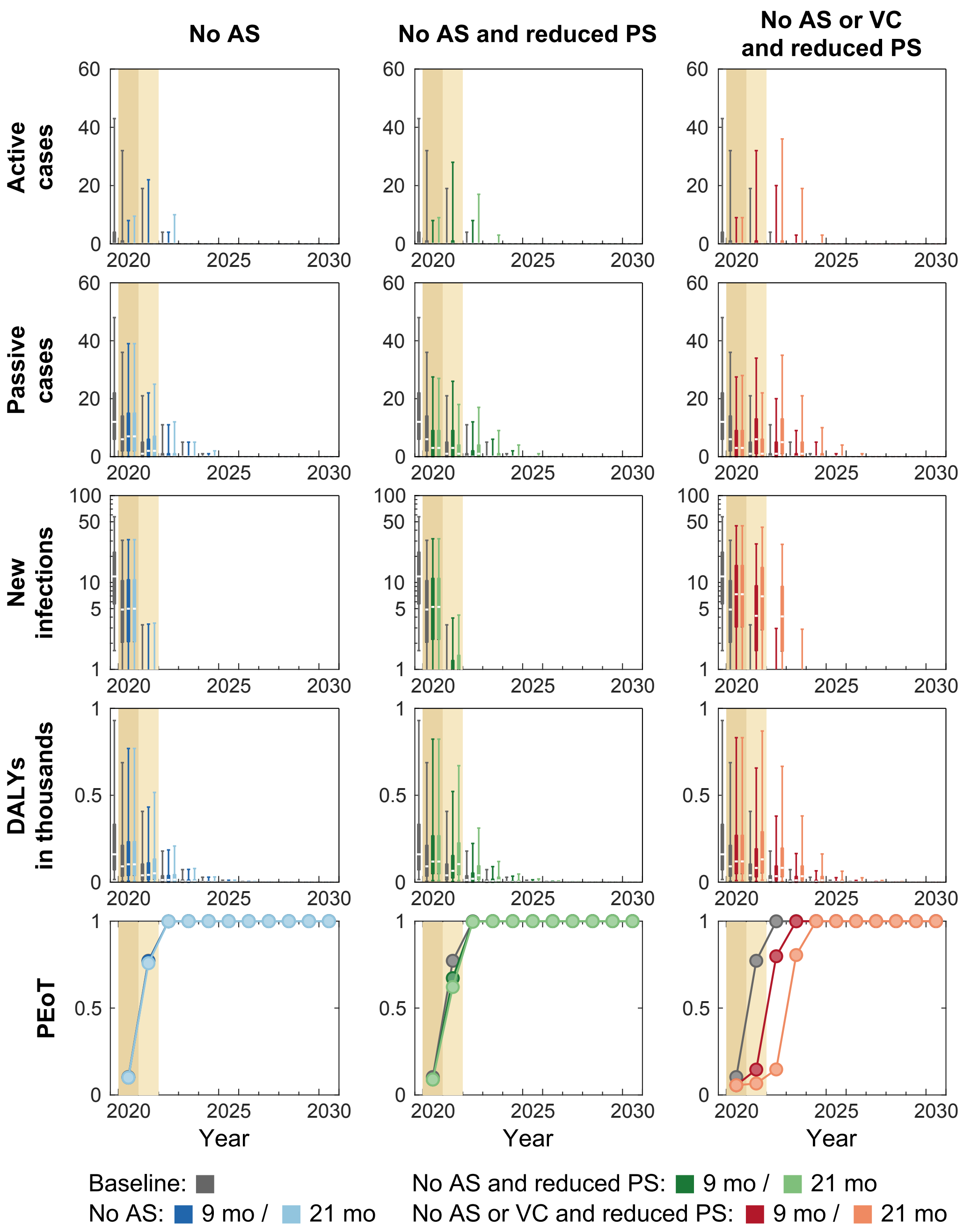


**Figure F. Time series of model outputs in Bolobo health zone under the baseline and six interruption scenarios.** During 2014–2018, an average of 48% of the population participated in active screening resulting in 9.97 reported cases (from both active and passive screenings) per 10,000 per annum in Bolobo health zone. The first Tiny Target deployment of vector control was scheduled in mid-2020. Interruptions by COVID-19 are assumed to take place in April 2020 and last until the end of 2020 or 2021 in our simulations. There are n = 10,000 independent samples, 10 from each of 1,000 independent samples from the joint posterior distributions of the fitted model parameters. Box plots summarise parameter and observational uncertainty. The lines in the boxes represent the medians of predicted results. The lower and upper bounds of the boxes indicate 25th and 75th percentiles. The minimum and maximum values are 2.5th and 97.5th percentiles and therefore whiskers cover 95% prediction intervals.

AS: active screening; PS: passive screening; VC: vector control; DALYs: disability-adjusted life years; PEoT: probability of elimination of transmission

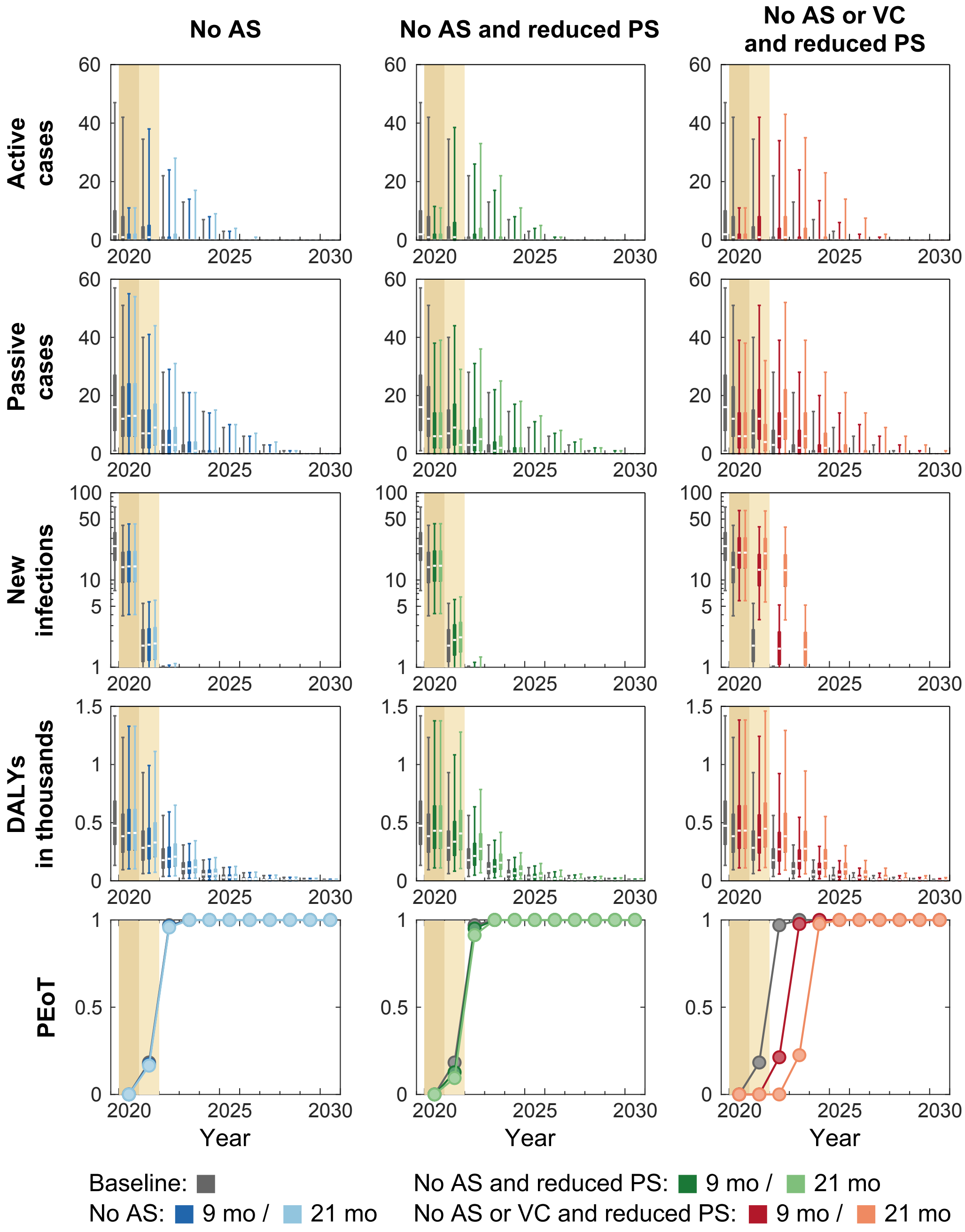


**Figure G. Time series of model outputs in Bulungu health zone under the baseline and six interruption scenarios.** During 2014–2018, an average of 17% of the population participated in active screening resulting in 2.05 reported cases (from both active and passive screenings) per 10,000 per annum in Bulungu health zone. The first Tiny Target deployment of vector control was scheduled in mid-2020. Interruptions by COVID-19 are assumed to take place in April 2020 and last until the end of 2020 or 2021 in our simulations. There are n = 10,000 independent samples, 10 from each of 1,000 independent samples from the joint posterior distributions of the fitted model parameters. Box plots summarise parameter and observational uncertainty. The lines in the boxes represent the medians of predicted results. The lower and upper bounds of the boxes indicate 25th and 75th percentiles. The minimum and maximum values are 2.5th and 97.5th percentiles and therefore whiskers cover 95% prediction intervals.
AS: active screening; PS: passive screening; VC: vector control; DALYs: disability-adjusted life years; PEoT: probability of elimination of transmission

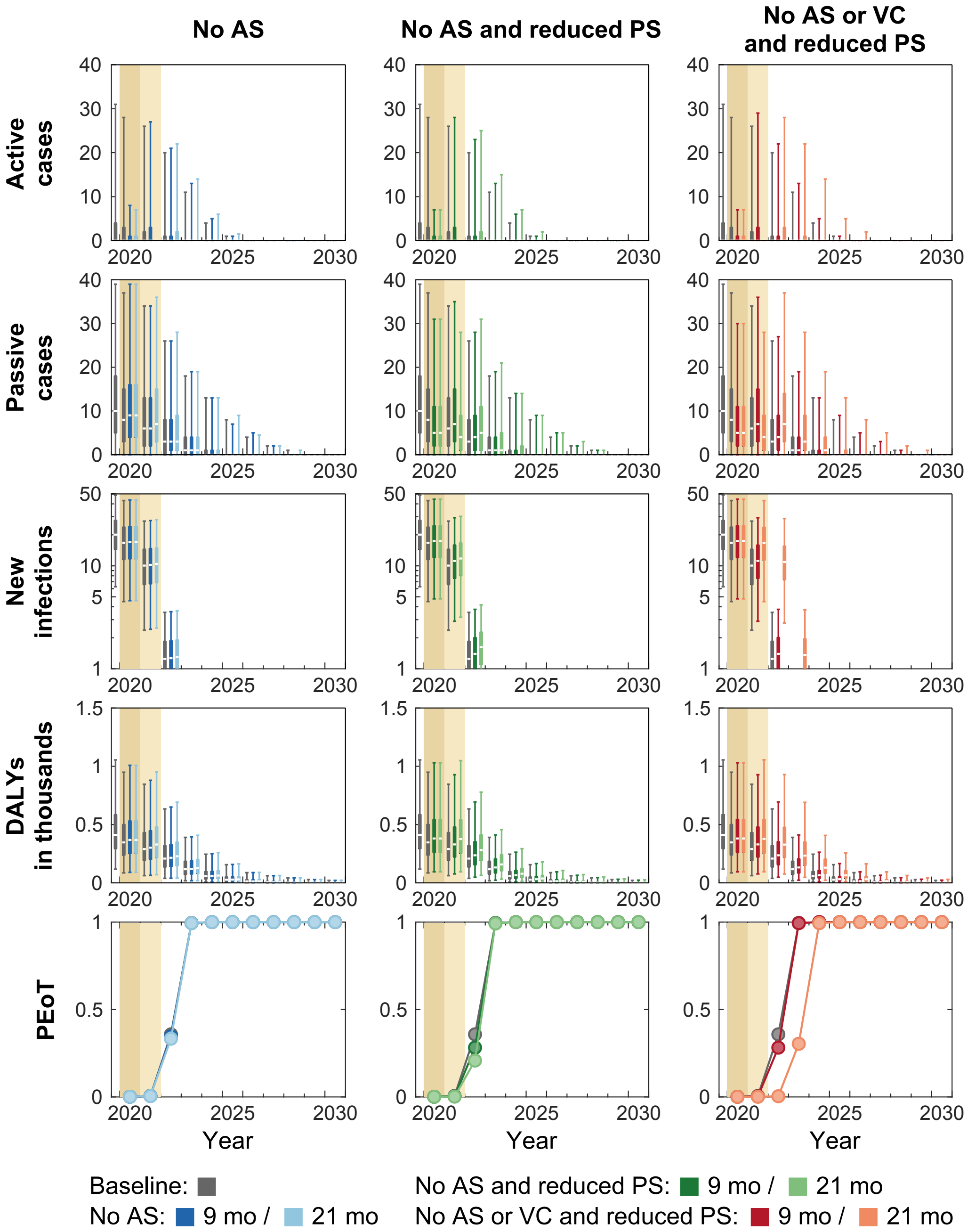


**Figure H. Time series of model outputs in Mokala health zone under the baseline and six interruption scenarios.** During 2014–2018, an average of 20% of the population participated in active screening resulting in 2.18 reported cases (from both active and passive screenings) per 10,000 per annum in Mokala health zone. The first Tiny Target deployment of vector control was scheduled in mid-2021. Interruptions by COVID-19 are assumed to take place in April 2020 and last until the end of 2020 or 2021 in our simulations. There are n = 10,000 independent samples, 10 from each of 1,000 independent samples from the joint posterior distributions of the fitted model parameters. Box plots summarise parameter and observational uncertainty. The lines in the boxes represent the medians of predicted results. The lower and upper bounds of the boxes indicate 25th and 75th percentiles. The minimum and maximum values are 2.5th and 97.5th percentiles and therefore whiskers cover 95% prediction intervals.

AS: active screening; PS: passive screening; VC: vector control; DALYs: disability-adjusted life years; PEoT: probability of elimination of transmission

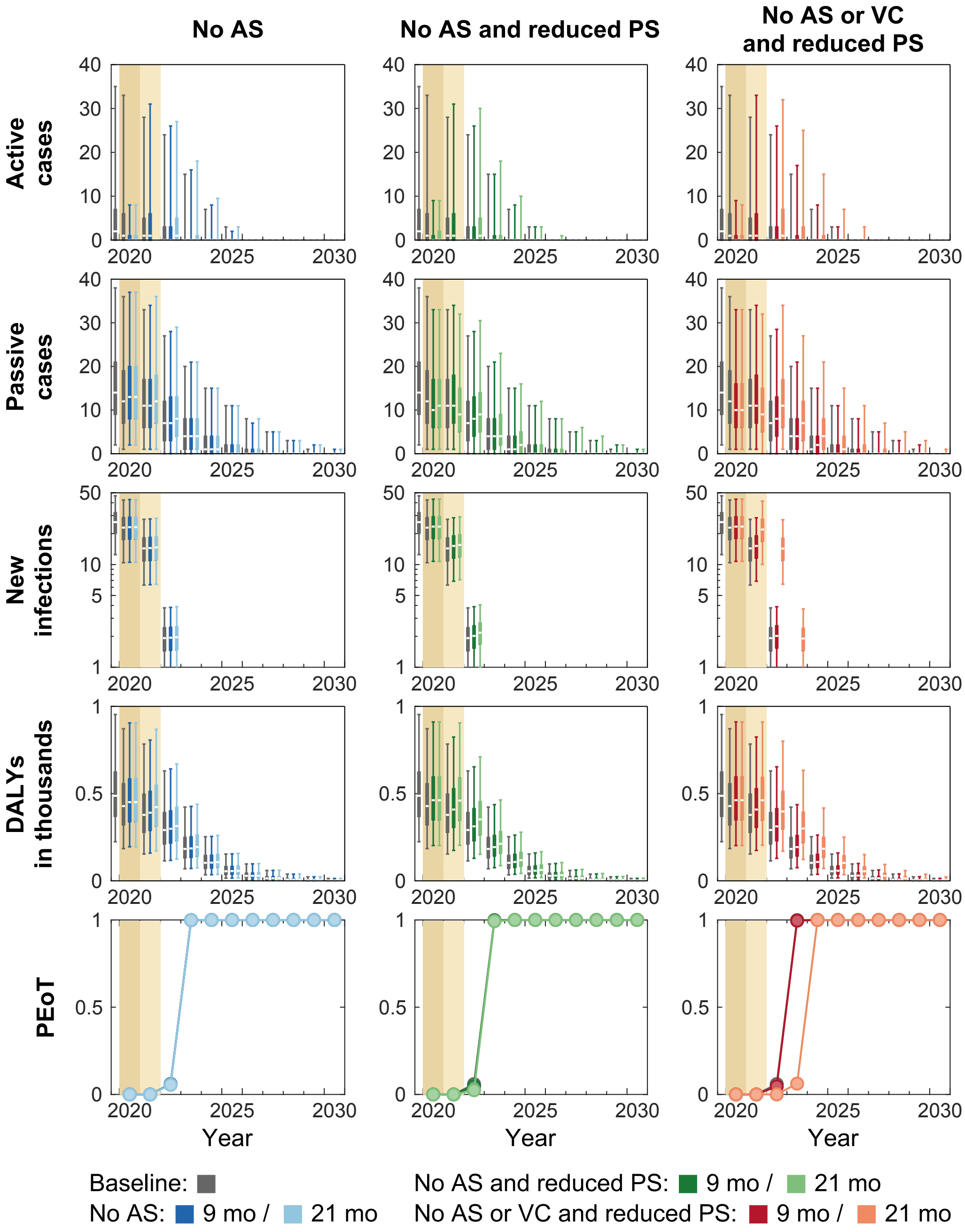


**Figure I. Time series of model outputs in Mushie health zone under the baseline and six interruption scenarios.** During 2014–2018, an average of 29% of the population participated in active screening resulting in 3.83 reported cases (from both active and passive screenings) per 10,000 per annum in Mushie health zone. The first Tiny Target deployment of vector control was scheduled in mid-2021. Interruptions by COVID-19 are assumed to take place in April 2020 and last until the end of 2020 or 2021 in our simulations. There are n = 10,000 independent samples, 10 from each of 1,000 independent samples from the joint posterior distributions of the fitted model parameters. Box plots summarise parameter and observational uncertainty. The lines in the boxes represent the medians of predicted results. The lower and upper bounds of the boxes indicate 25th and 75th percentiles. The minimum and maximum values are 2.5th and 97.5th percentiles and therefore whiskers cover 95% prediction intervals.

AS: active screening; PS: passive screening; VC: vector control; DALYs: disability-adjusted life years; PEoT: probability of elimination of transmission

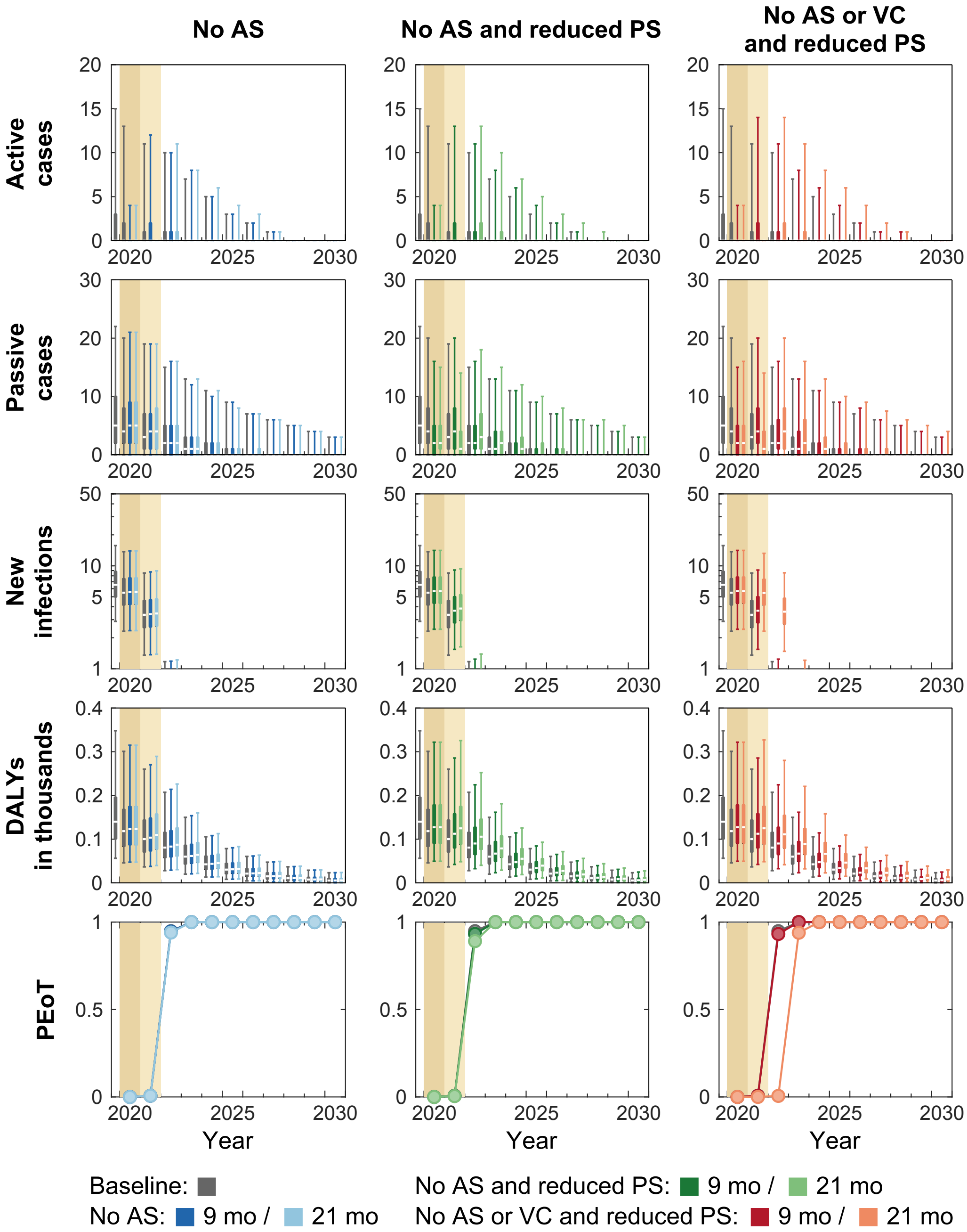


**Figure J. Time series of model outputs in Yumbi health zone under the baseline and six interruption scenarios.** During 2014–2018, an average of 15% of the population participated in active screening resulting in 1.94 reported cases (from both active and passive screenings) per 10,000 per annum in Yumbi health zone. The first Tiny Target deployment of vector control was scheduled in mid-2021. Interruptions by COVID-19 are assumed to take place in April 2020 and last until the end of 2020 or 2021 in our simulations. There are n = 10,000 independent samples, 10 from each of 1,000 independent samples from the joint posterior distributions of the fitted model parameters. Box plots summarise parameter and observational uncertainty. The lines in the boxes represent the medians of predicted results. The lower and upper bounds of the boxes indicate 25th and 75th percentiles. The minimum and maximum values are 2.5th and 97.5th percentiles and therefore whiskers cover 95% prediction intervals.

AS: active screening; PS: passive screening; VC: vector control; DALYs: disability-adjusted life years; PEoT: probability of elimination of transmission

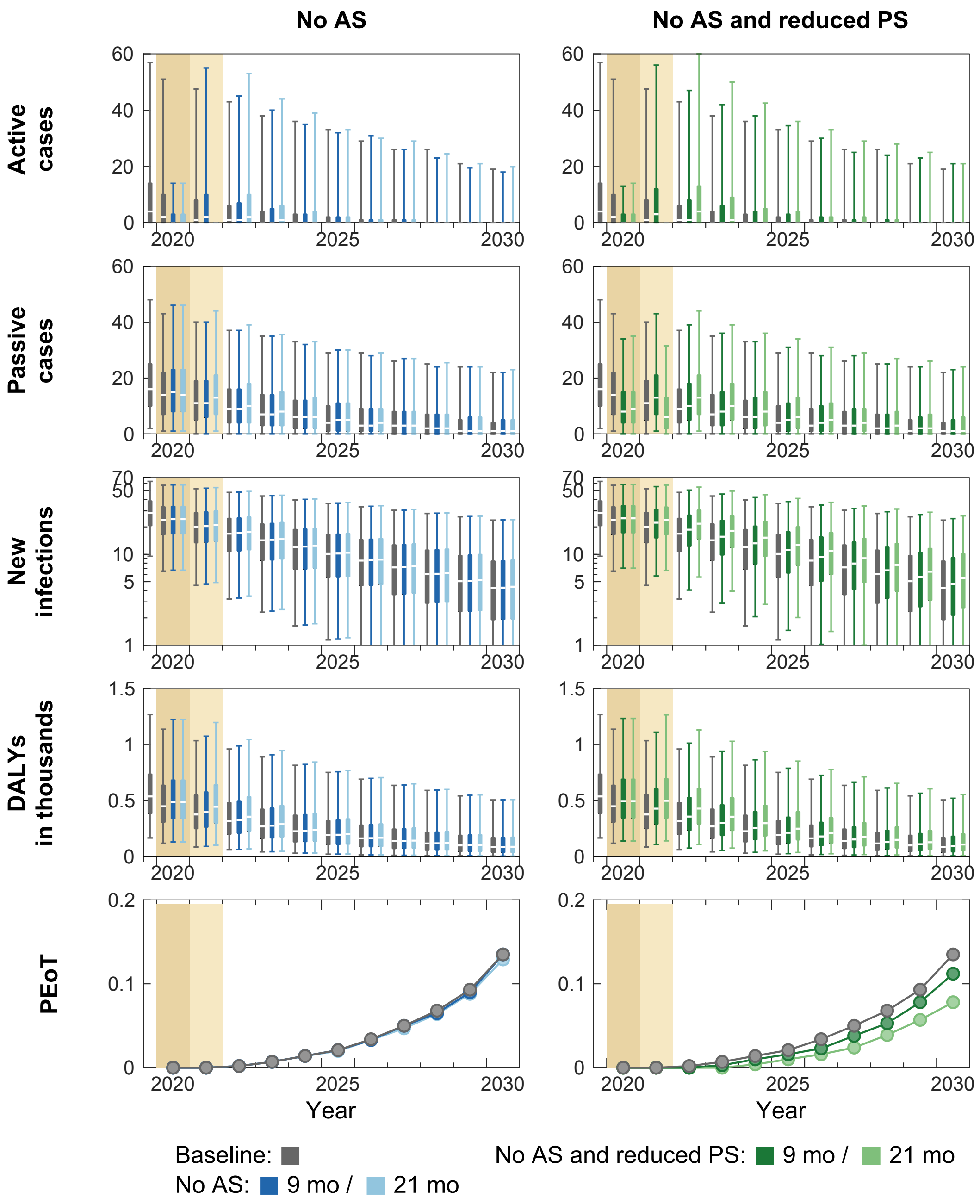


**Figure K. Time series of model outputs in Bagata health zone under the baseline and six interruption scenarios.** During 2014–2018, an average of 35% of the population participated in active screening resulting in 3.28 reported cases (from both active and passive screenings) per 10,000 per annum in Bagata health zone. There are n = 10,000 independent samples, 10 from each of 1,000 independent samples from the joint posterior distributions of the fitted model parameters. Box plots summarise parameter and observational uncertainty. The lines in the boxes represent the medians of predicted results. The lower and upper bounds of the boxes indicate 25th and 75th percentiles. The minimum and maximum values are 2.5th and 97.5th percentiles and therefore whiskers cover 95% prediction intervals.

AS: active screening; PS: passive screening; VC: vector control; DALYs: disability-adjusted life years; PEoT: probability of elimination of transmission

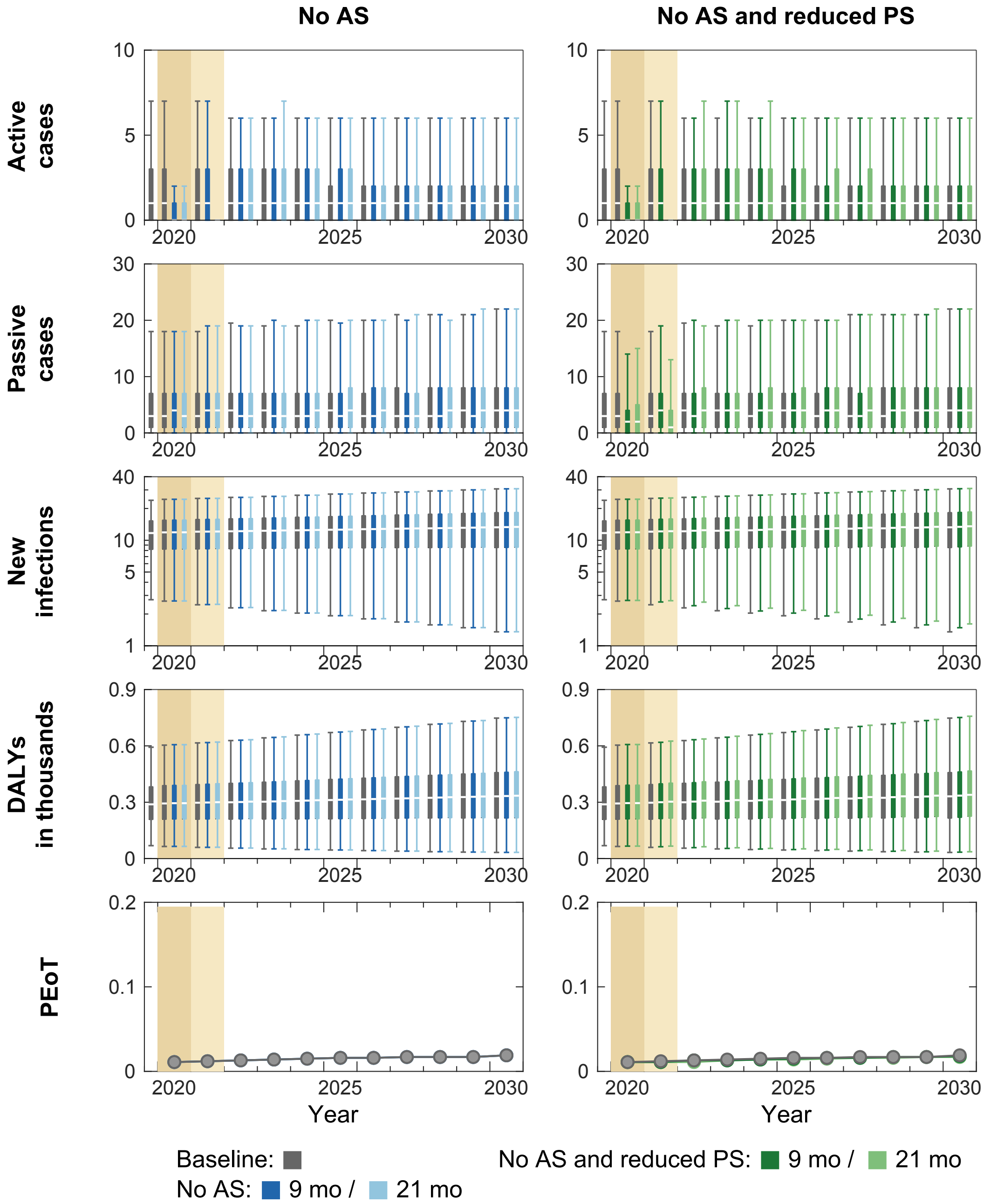


**Figure L. Time series of model outputs in Bandjau health zone under the baseline and six interruption scenarios.** During 2014–2018, an average of 1% of the population participated in active screening resulting in 0.43 reported cases (from both active and passive screenings) per 10,000 per annum in Bandjau health zone. There are n = 10,000 independent samples, 10 from each of 1,000 independent samples from the joint posterior distributions of the fitted model parameters. Box plots summarise parameter and observational uncertainty. The lines in the boxes represent the medians of predicted results. The lower and upper bounds of the boxes indicate 25th and 75th percentiles. The minimum and maximum values are 2.5th and 97.5th percentiles and therefore whiskers cover 95% prediction intervals.
AS: active screening; PS: passive screening; VC: vector control; DALYs: disability-adjusted life years; PEoT: probability of elimination of transmission

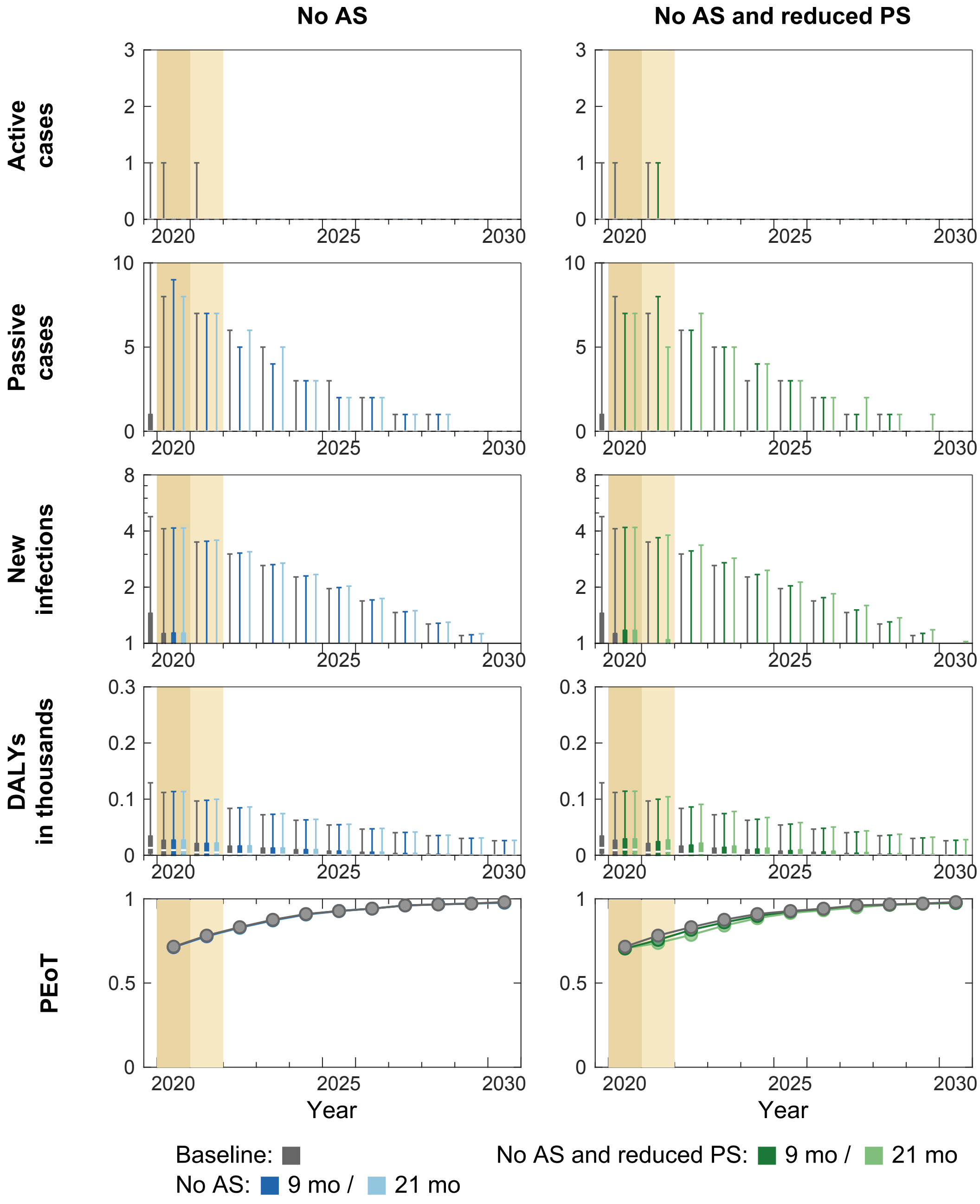


**Figure M. Time series of model outputs in Boko health zone under the baseline and six interruption scenarios.** During 2014–2018, an average of 3% of the population participated in active screening resulting in 0.14 reported cases (from both active and passive screenings) per 10,000 per annum in Boko health zone. There are n = 10,000 independent samples, 10 from each of 1,000 independent samples from the joint posterior distributions of the fitted model parameters. Box plots summarise parameter and observational uncertainty. The lines in the boxes represent the medians of predicted results. The lower and upper bounds of the boxes indicate 25th and 75th percentiles. The minimum and maximum values are 2.5th and 97.5th percentiles and therefore whiskers cover 95% prediction intervals.

AS: active screening; PS: passive screening; VC: vector control; DALYs: disability-adjusted life years; PEoT: probability of elimination of transmission

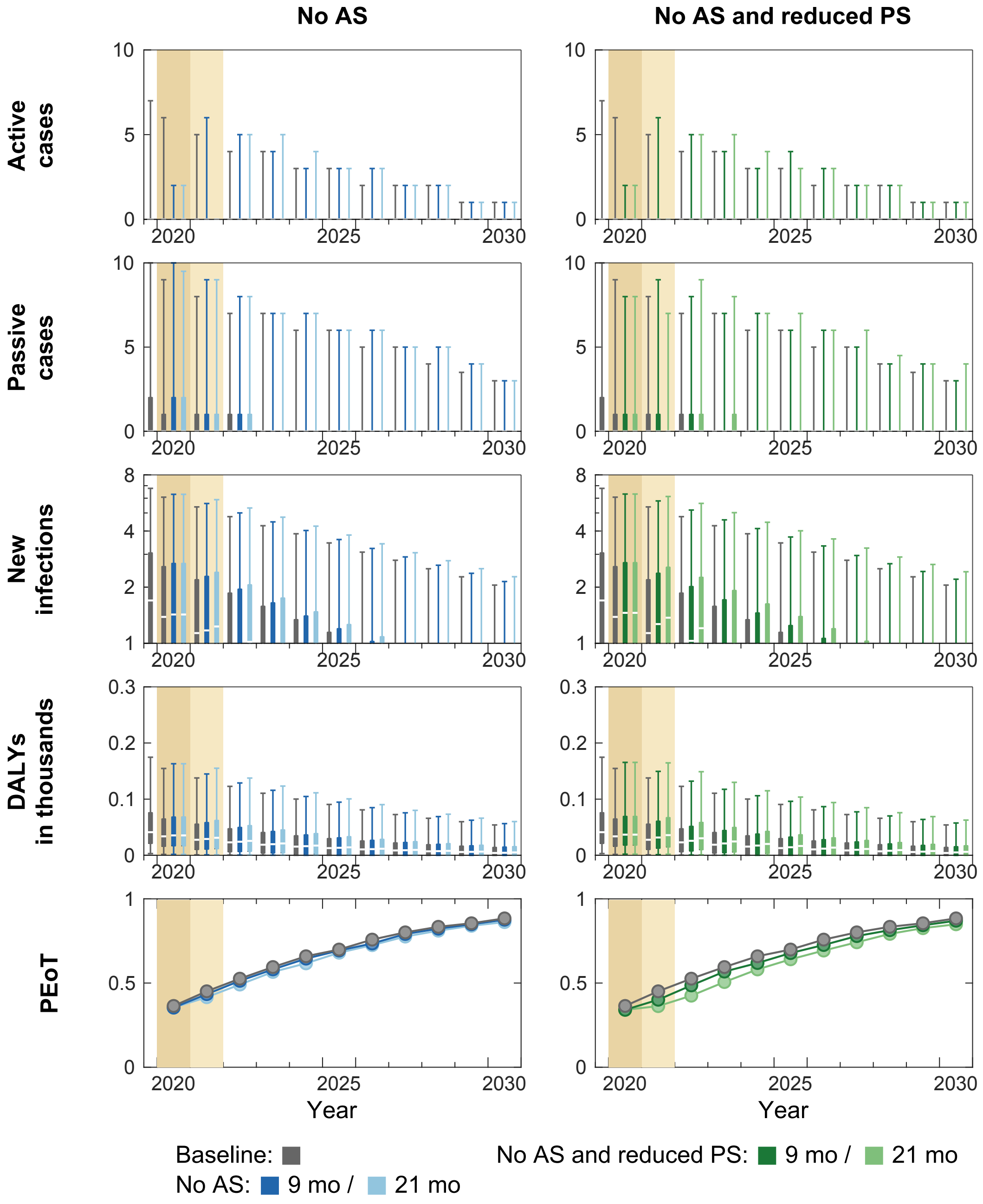


**Figure N. Time series of model outputs in Bosobe health zone under the baseline and six interruption scenarios.** During 2014–2018, an average of 9% of the population participated in active screening resulting in 0.4 reported cases (from both active and passive screenings) per 10,000 per annum in Bosobe health zone. There are n = 10,000 independent samples, 10 from each of 1,000 independent samples from the joint posterior distributions of the fitted model parameters. Box plots summarise parameter and observational uncertainty. The lines in the boxes represent the medians of predicted results. The lower and upper bounds of the boxes indicate 25th and 75th percentiles. The minimum and maximum values are 2.5th and 97.5th percentiles and therefore whiskers cover 95% prediction intervals.
AS: active screening; PS: passive screening; VC: vector control; DALYs: disability-adjusted life years; PEoT: probability of elimination of transmission

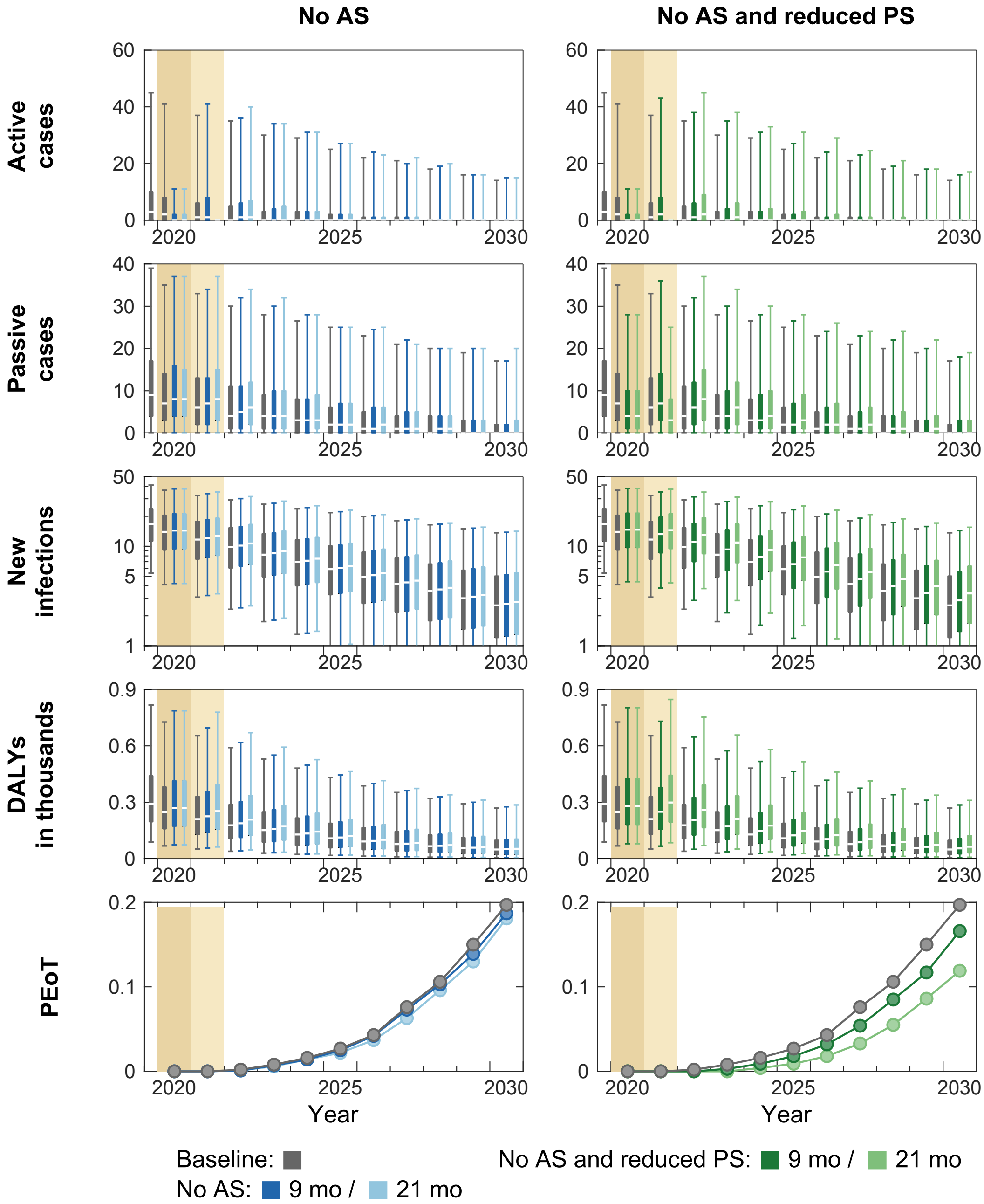


**Figure O. Time series of model outputs in Djuma health zone under the baseline and six interruption scenarios.** During 2014–2018, an average of 21% of the population participated in active screening resulting in 0.96 reported cases (from both active and passive screenings) per 10,000 per annum in Djuma health zone. There are n = 10,000 independent samples, 10 from each of 1,000 independent samples from the joint posterior distributions of the fitted model parameters. Box plots summarise parameter and observational uncertainty. The lines in the boxes represent the medians of predicted results. The lower and upper bounds of the boxes indicate 25th and 75th percentiles. The minimum and maximum values are 2.5th and 97.5th percentiles and therefore whiskers cover 95% prediction intervals.
AS: active screening; PS: passive screening; VC: vector control; DALYs: disability-adjusted life years; PEoT: probability of elimination of transmission

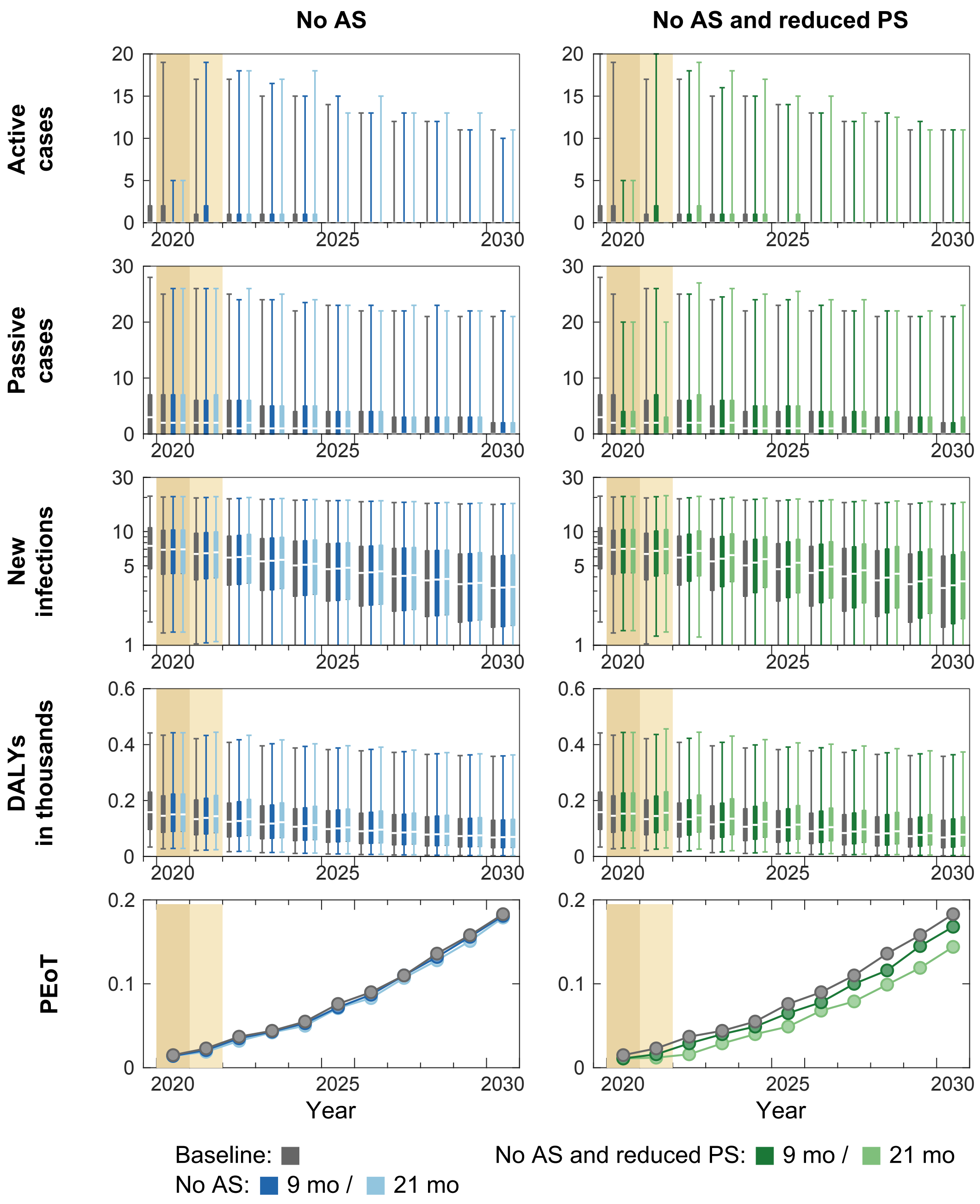


**Figure P. Time series of model outputs in Idiofa health zone under the baseline and six interruption scenarios.** During 2014–2018, an average of 11% of the population participated in active screening resulting in 0.99 reported cases (from both active and passive screenings) per 10,000 per annum in Idiofa health zone. There are n = 10,000 independent samples, 10 from each of 1,000 independent samples from the joint posterior distributions of the fitted model parameters. Box plots summarise parameter and observational uncertainty. The lines in the boxes represent the medians of predicted results. The lower and upper bounds of the boxes indicate 25th and 75th percentiles. The minimum and maximum values are 2.5th and 97.5th percentiles and therefore whiskers cover 95% prediction intervals.

AS: active screening; PS: passive screening; VC: vector control; DALYs: disability-adjusted life years; PEoT: probability of elimination of transmission

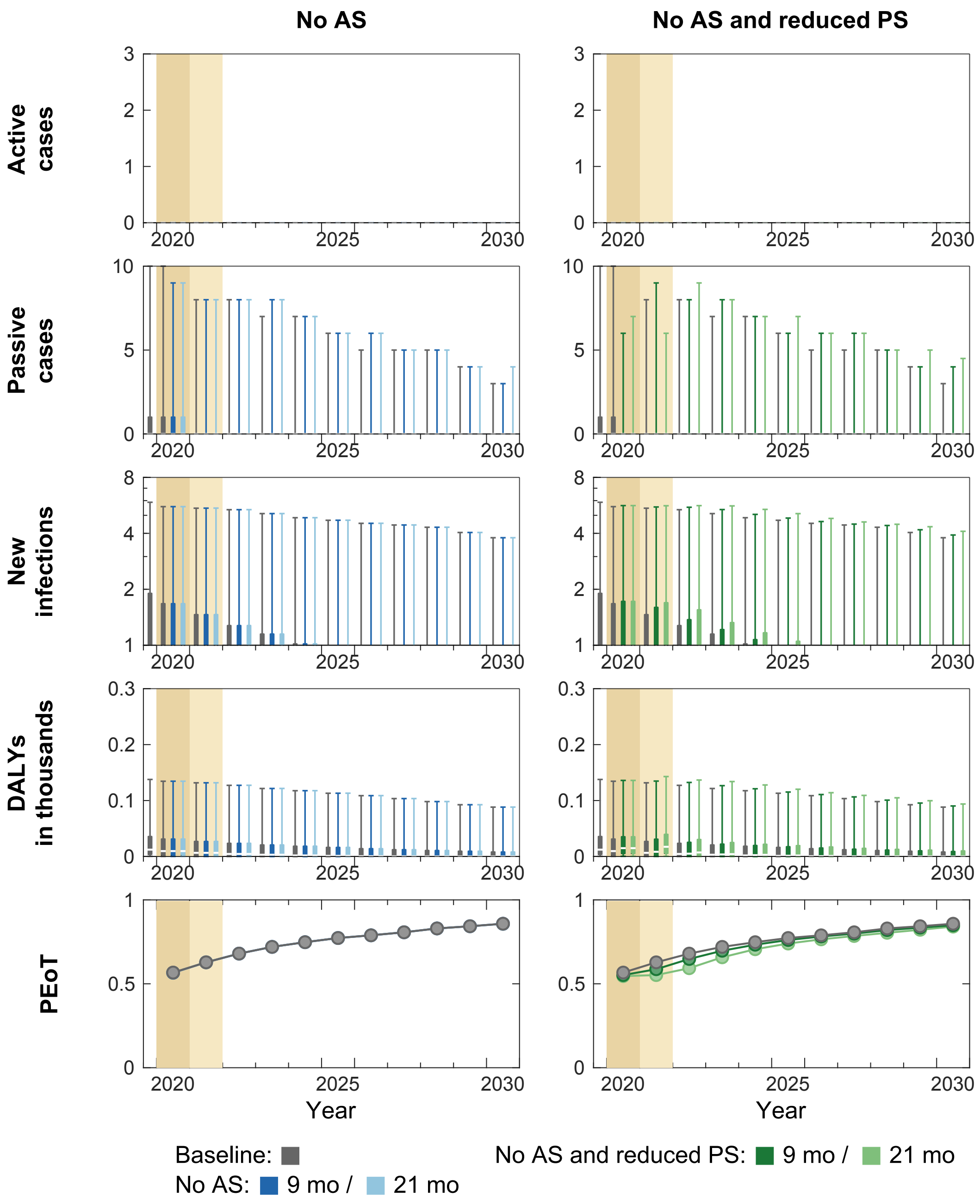


**Figure Q. Time series of model outputs in Inongo health zone under the baseline and six interruption scenarios.** During 2014–2018, an average of 0% of the population participated in active screening resulting in 0.08 reported cases (from both active and passive screenings) per 10,000 per annum in Inongo health zone. There are n = 10,000 independent samples, 10 from each of 1,000 independent samples from the joint posterior distributions of the fitted model parameters. Box plots summarise parameter and observational uncertainty. The lines in the boxes represent the medians of predicted results. The lower and upper bounds of the boxes indicate 25th and 75th percentiles. The minimum and maximum values are 2.5th and 97.5th percentiles and therefore whiskers cover 95% prediction intervals.
AS: active screening; PS: passive screening; VC: vector control; DALYs: disability-adjusted life years; PEoT: probability of elimination of transmission

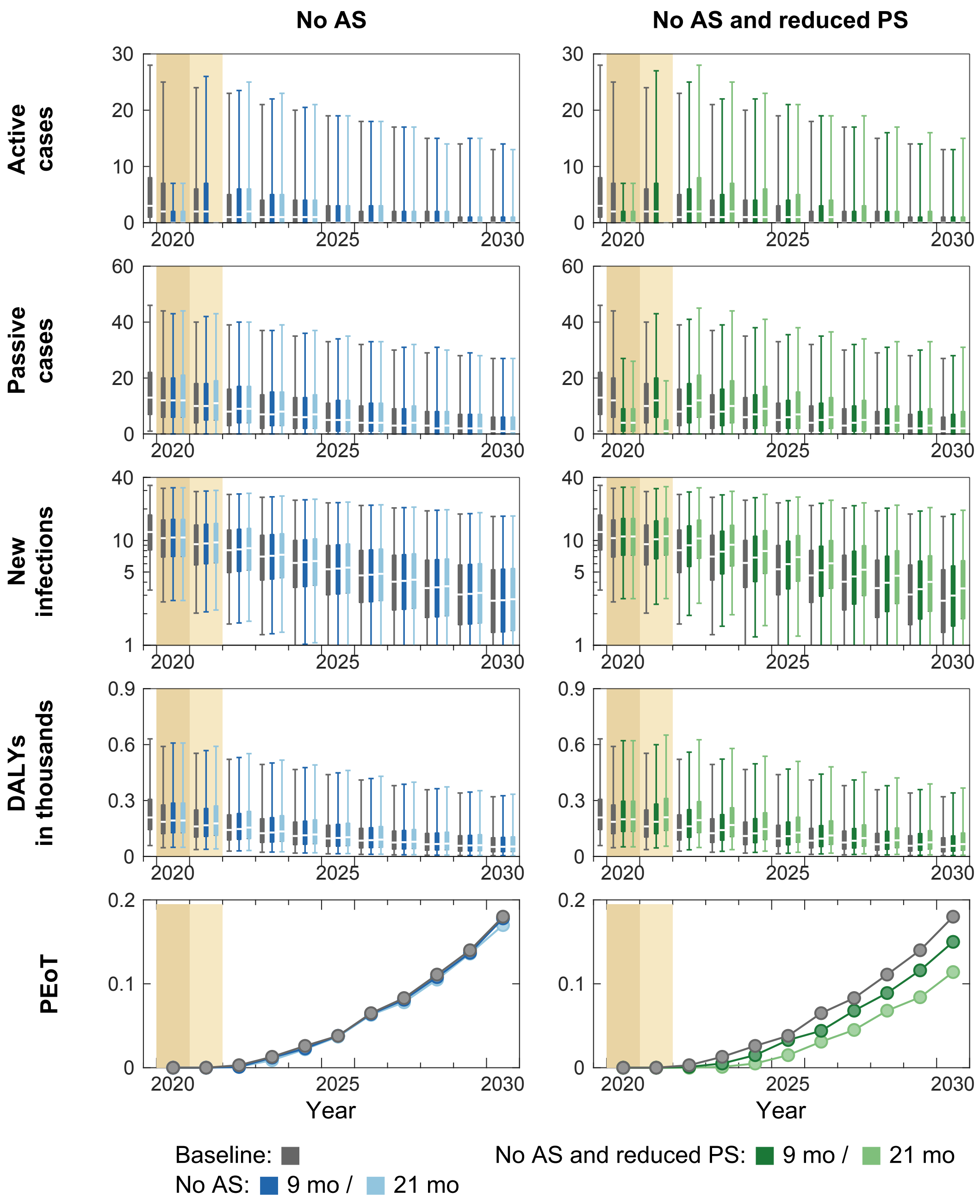


**Figure R. Time series of model outputs in Ipamu health zone under the baseline and six interruption scenarios.** During 2014–2018, an average of 10% of the population participated in active screening resulting in 1.52 reported cases (from both active and passive screenings) per 10,000 per annum in Ipamu health zone. There are n = 10,000 independent samples, 10 from each of 1,000 independent samples from the joint posterior distributions of the fitted model parameters. Box plots summarise parameter and observational uncertainty. The lines in the boxes represent the medians of predicted results. The lower and upper bounds of the boxes indicate 25th and 75th percentiles. The minimum and maximum values are 2.5th and 97.5th percentiles and therefore whiskers cover 95% prediction intervals.
AS: active screening; PS: passive screening; VC: vector control; DALYs: disability-adjusted life years; PEoT: probability of elimination of transmission

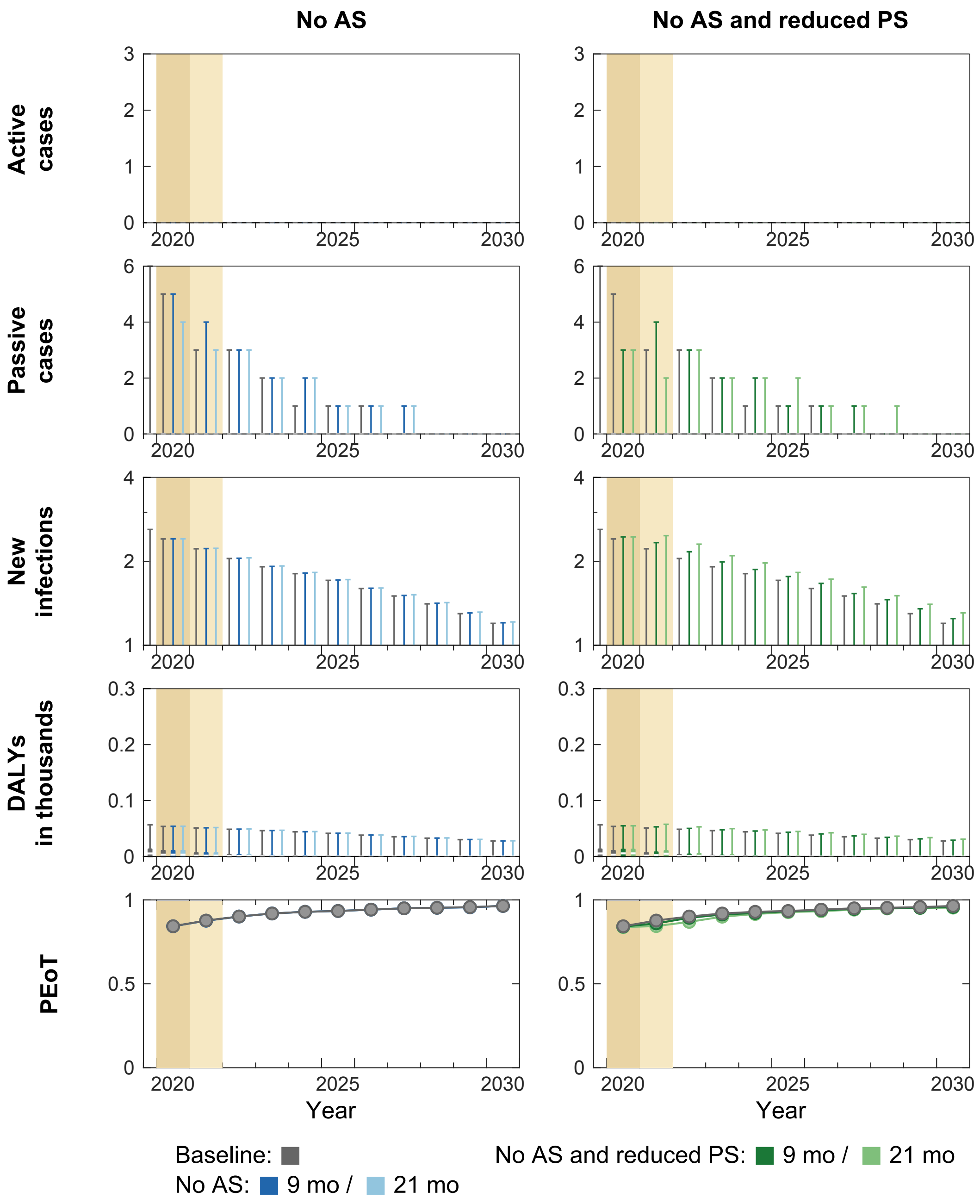


**Figure S. Time series of model outputs in Kasongolunda health zone under the baseline and six interruption scenarios.** During 2014–2018, an average of 2% of the population participated in active screening resulting in 0.06 reported cases (from both active and passive screenings) per 10,000 per annum in Kasongolunda health zone. There are n = 10,000 independent samples, 10 from each of 1,000 independent samples from the joint posterior distributions of the fitted model parameters. Box plots summarise parameter and observational uncertainty. The lines in the boxes represent the medians of predicted results. The lower and upper bounds of the boxes indicate 25th and 75th percentiles. The minimum and maximum values are 2.5th and 97.5th percentiles and therefore whiskers cover 95% prediction intervals.

AS: active screening; PS: passive screening; VC: vector control; DALYs: disability-adjusted life years; PEoT: probability of elimination of transmission

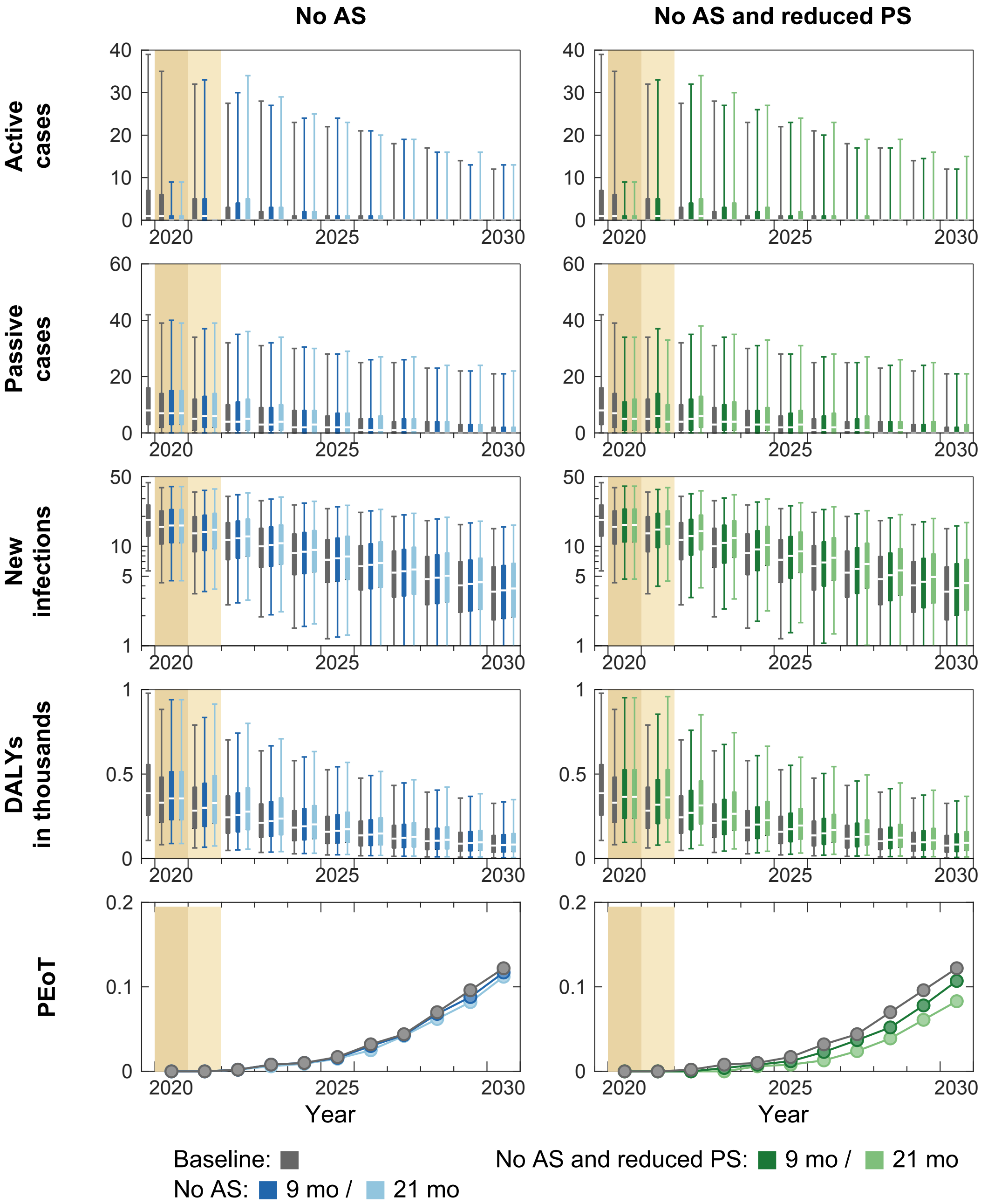


**Figure T. Time series of model outputs in Kenge health zone under the baseline and six interruption scenarios.** During 2014–2018, an average of 15% of the population participated in active screening resulting in 0.71 reported cases (from both active and passive screenings) per 10,000 per annum in Kenge health zone. There are n = 10,000 independent samples, 10 from each of 1,000 independent samples from the joint posterior distributions of the fitted model parameters. Box plots summarise parameter and observational uncertainty. The lines in the boxes represent the medians of predicted results. The lower and upper bounds of the boxes indicate 25th and 75th percentiles. The minimum and maximum values are 2.5th and 97.5th percentiles and therefore whiskers cover 95% prediction intervals.

AS: active screening; PS: passive screening; VC: vector control; DALYs: disability-adjusted life years; PEoT: probability of elimination of transmission

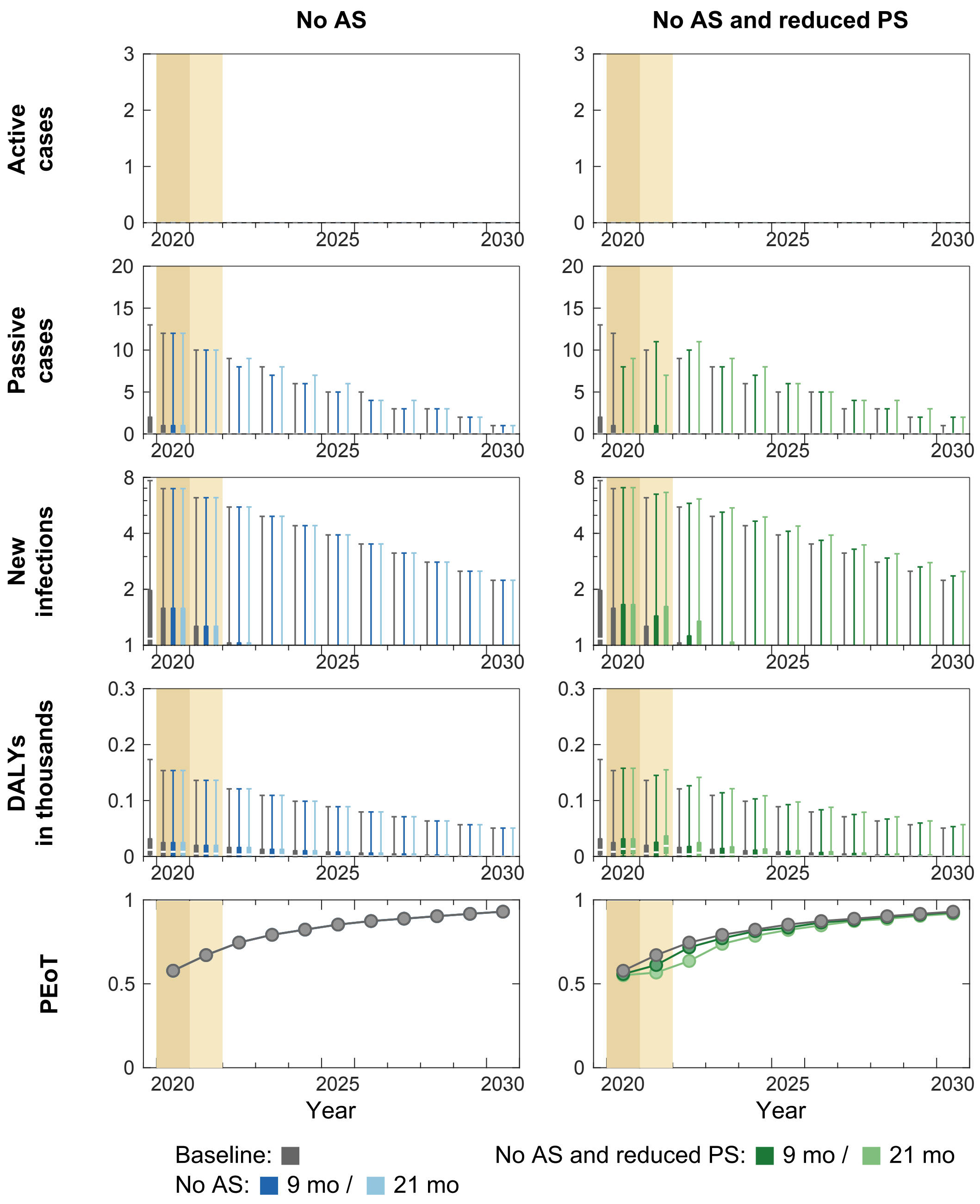


**Figure U. Time series of model outputs in Kikwit Nord health zone under the baseline and six interruption scenarios.** During 2014–2018, an average of 0% of the population participated in active screening resulting in 0.08 reported cases (from both active and passive screenings) per 10,000 per annum in Kikwit Nord health zone. There are n = 10,000 independent samples, 10 from each of 1,000 independent samples from the joint posterior distributions of the fitted model parameters. Box plots summarise parameter and observational uncertainty. The lines in the boxes represent the medians of predicted results. The lower and upper bounds of the boxes indicate 25th and 75th percentiles. The minimum and maximum values are 2.5th and 97.5th percentiles and therefore whiskers cover 95% prediction intervals.

AS: active screening; PS: passive screening; VC: vector control; DALYs: disability-adjusted life years; PEoT: probability of elimination of transmission

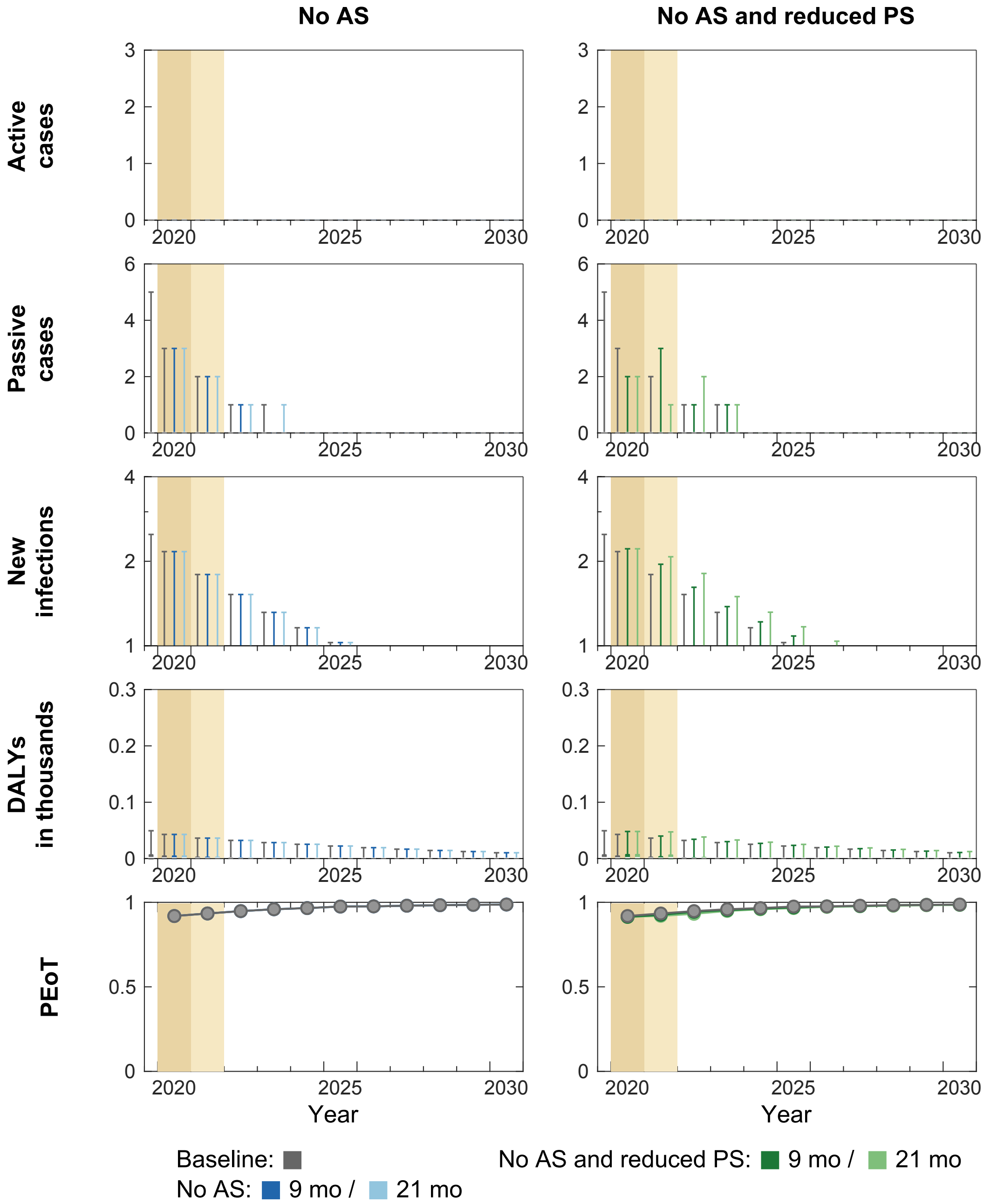


**Figure V. Time series of model outputs in Kikwit Sud health zone under the baseline and six interruption scenarios.** During 2014–2018, an average of 0% of the population participated in active screening resulting in 0.04 reported cases (from both active and passive screenings) per 10,000 per annum in Kikwit Sud health zone. There are n = 10,000 independent samples, 10 from each of 1,000 independent samples from the joint posterior distributions of the fitted model parameters. Box plots summarise parameter and observational uncertainty. The lines in the boxes represent the medians of predicted results. The lower and upper bounds of the boxes indicate 25th and 75th percentiles. The minimum and maximum values are 2.5th and 97.5th percentiles and therefore whiskers cover 95% prediction intervals.

AS: active screening; PS: passive screening; VC: vector control; DALYs: disability-adjusted life years; PEoT: probability of elimination of transmission

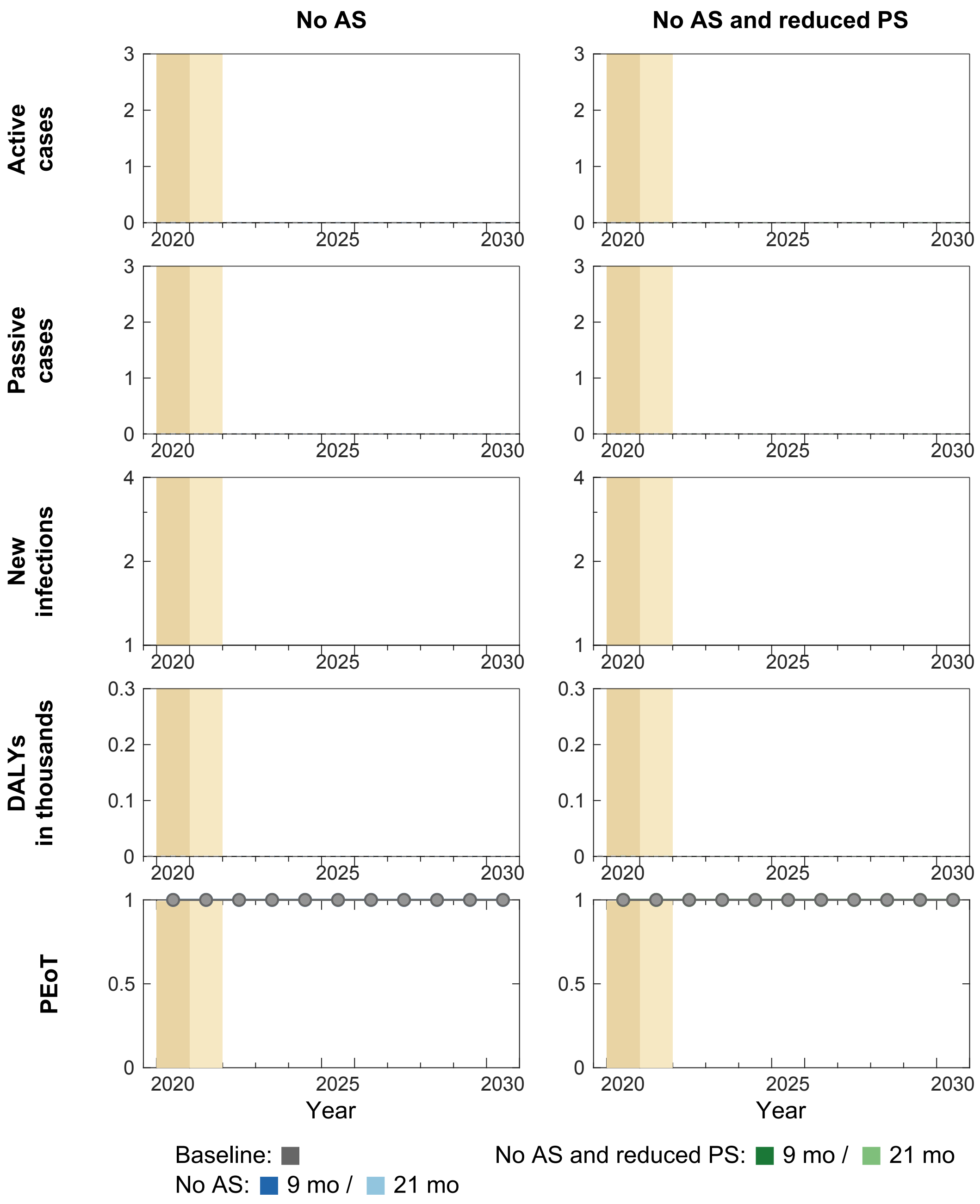


**Figure W. Time series of model outputs in Kimbau health zone under the baseline and six interruption scenarios.** During 2014–2018, an average of 1% of the population participated in active screening resulting in 0.01 reported cases (from both active and passive screenings) per 10,000 per annum in Kimbau health zone. There are n = 10,000 independent samples, 10 from each of 1,000 independent samples from the joint posterior distributions of the fitted model parameters. Box plots summarise parameter and observational uncertainty. The lines in the boxes represent the medians of predicted results. The lower and upper bounds of the boxes indicate 25th and 75th percentiles. The minimum and maximum values are 2.5th and 97.5th percentiles and therefore whiskers cover 95% prediction intervals.
AS: active screening; PS: passive screening; VC: vector control; DALYs: disability-adjusted life years; PEoT: probability of elimination of transmission

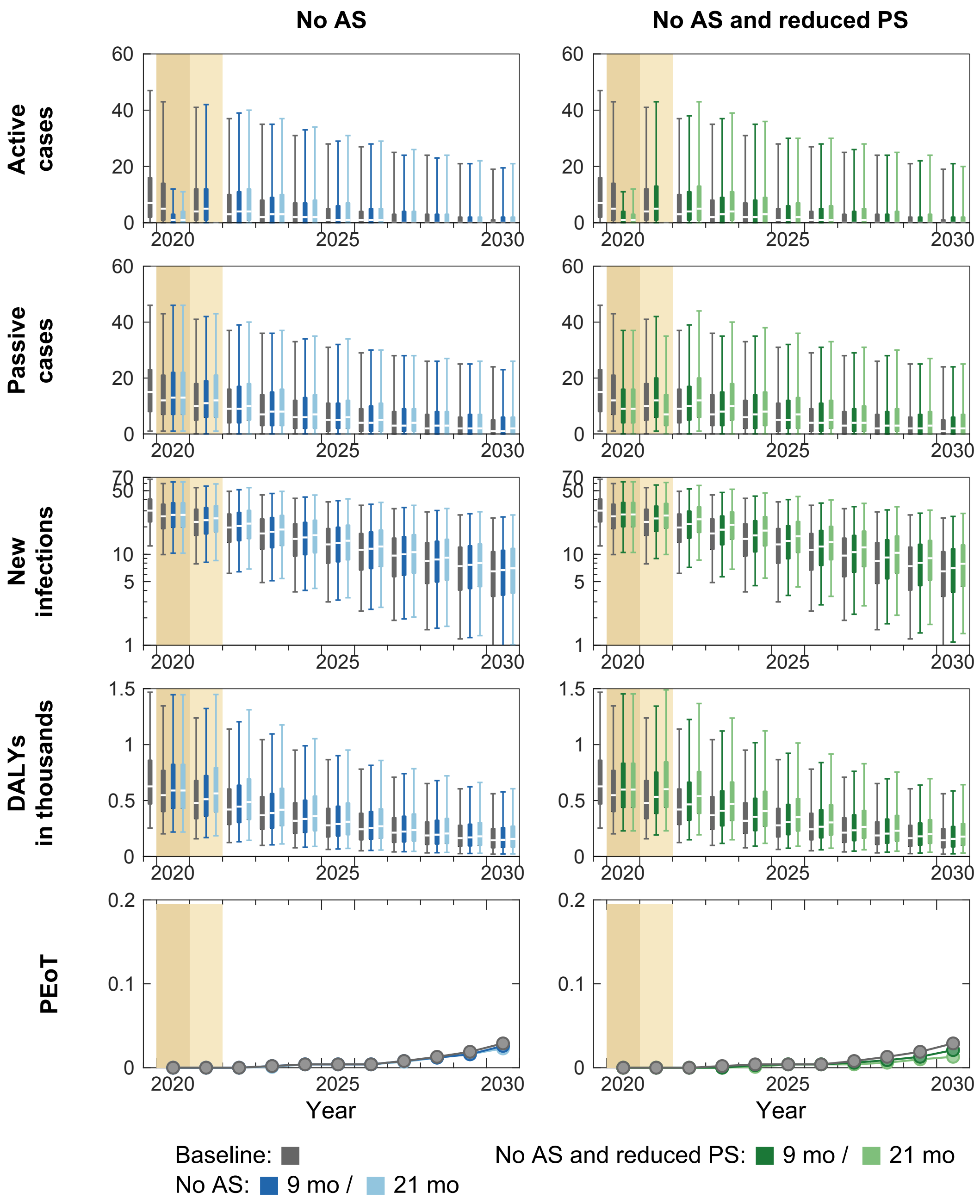


**Figure X. Time series of model outputs in Kimputu health zone under the baseline and six interruption scenarios.** During 2014–2018, an average of 16% of the population participated in active screening resulting in 2.16 reported cases (from both active and passive screenings) per 10,000 per annum in Kimputu health zone. There are n = 10,000 independent samples, 10 from each of 1,000 independent samples from the joint posterior distributions of the fitted model parameters. Box plots summarise parameter and observational uncertainty. The lines in the boxes represent the medians of predicted results. The lower and upper bounds of the boxes indicate 25th and 75th percentiles. The minimum and maximum values are 2.5th and 97.5th percentiles and therefore whiskers cover 95% prediction intervals.

AS: active screening; PS: passive screening; VC: vector control; DALYs: disability-adjusted life years; PEoT: probability of elimination of transmission

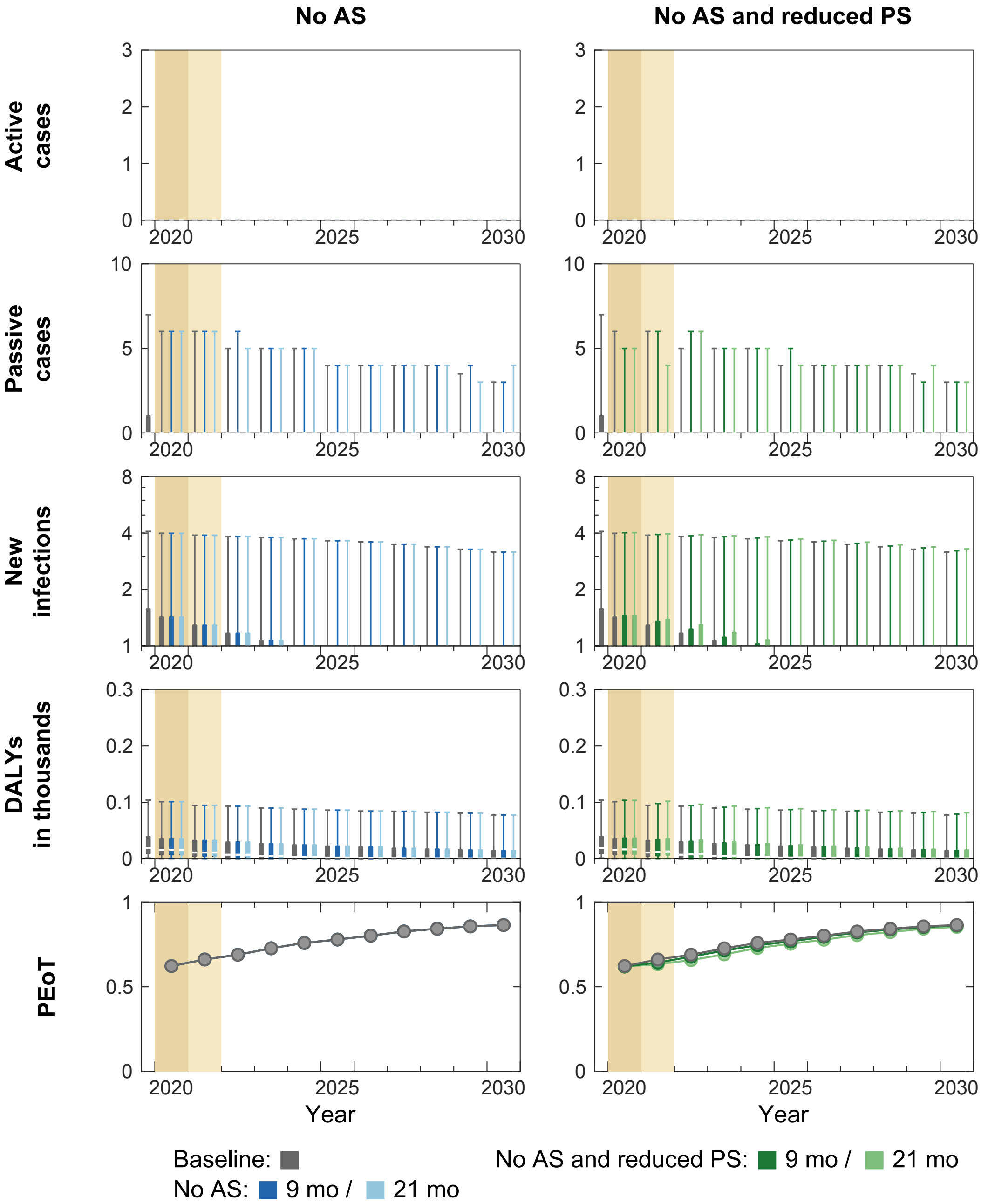


**Figure Y. Time series of model outputs in Kiri health zone under the baseline and six interruption scenarios.** During 2014–2018, an average of 0% of the population participated in active screening resulting in 0.05 reported cases (from both active and passive screenings) per 10,000 per annum in Kiri health zone. There are n = 10,000 independent samples, 10 from each of 1,000 independent samples from the joint posterior distributions of the fitted model parameters. Box plots summarise parameter and observational uncertainty. The lines in the boxes represent the medians of predicted results. The lower and upper bounds of the boxes indicate 25th and 75th percentiles. The minimum and maximum values are 2.5th and 97.5th percentiles and therefore whiskers cover 95% prediction intervals.

AS: active screening; PS: passive screening; VC: vector control; DALYs: disability-adjusted life years; PEoT: probability of elimination of transmission

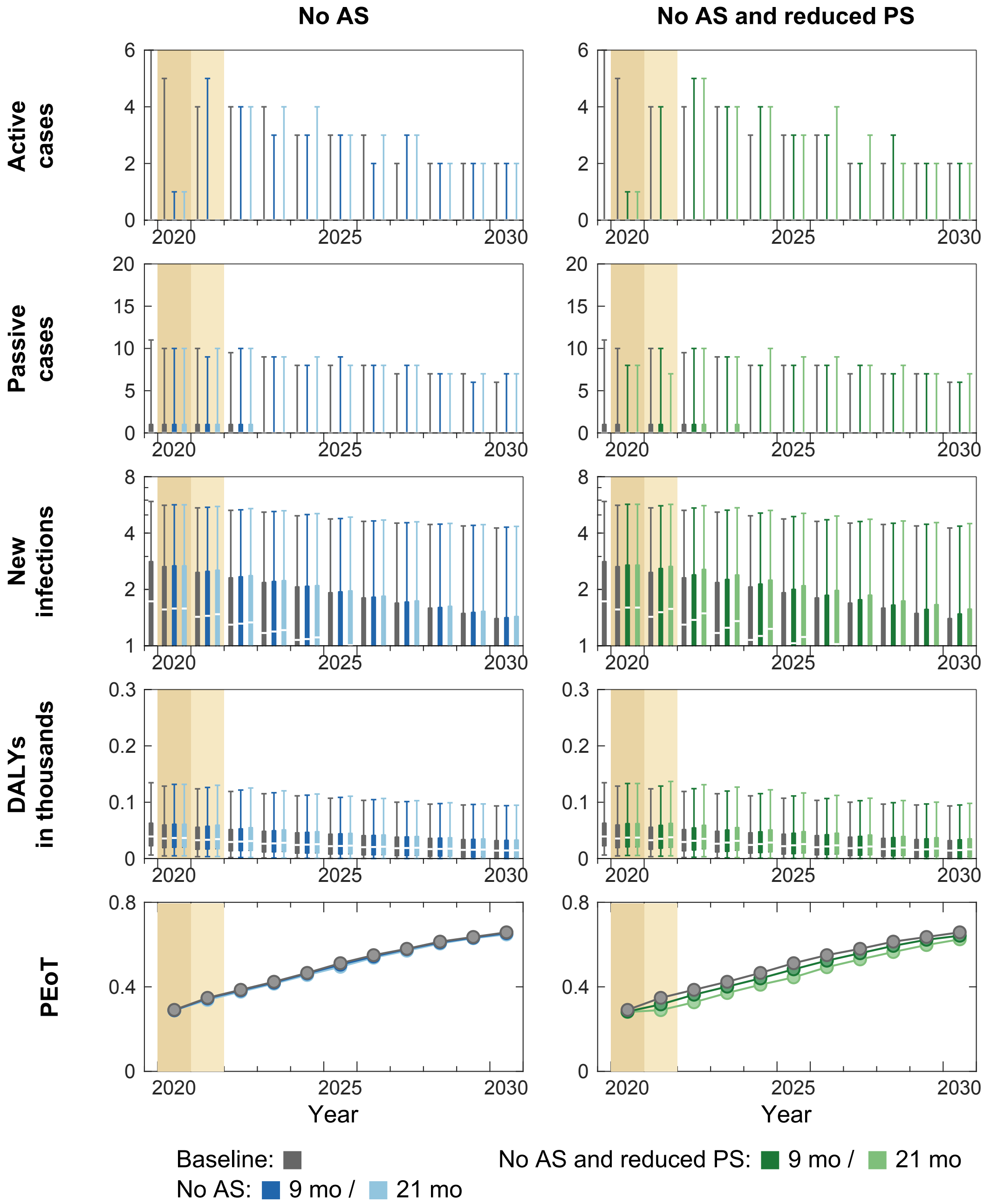


**Figure Z. Time series of model outputs in Koshibanda health zone under the baseline and six interruption scenarios.** During 2014–2018, an average of 8% of the population participated in active screening resulting in 0.5 reported cases (from both active and passive screenings) per 10,000 per annum in Koshibanda health zone. There are n = 10,000 independent samples, 10 from each of 1,000 independent samples from the joint posterior distributions of the fitted model parameters. Box plots summarise parameter and observational uncertainty. The lines in the boxes represent the medians of predicted results. The lower and upper bounds of the boxes indicate 25th and 75th percentiles. The minimum and maximum values are 2.5th and 97.5th percentiles and therefore whiskers cover 95% prediction intervals.

AS: active screening; PS: passive screening; VC: vector control; DALYs: disability-adjusted life years; PEoT: probability of elimination of transmission

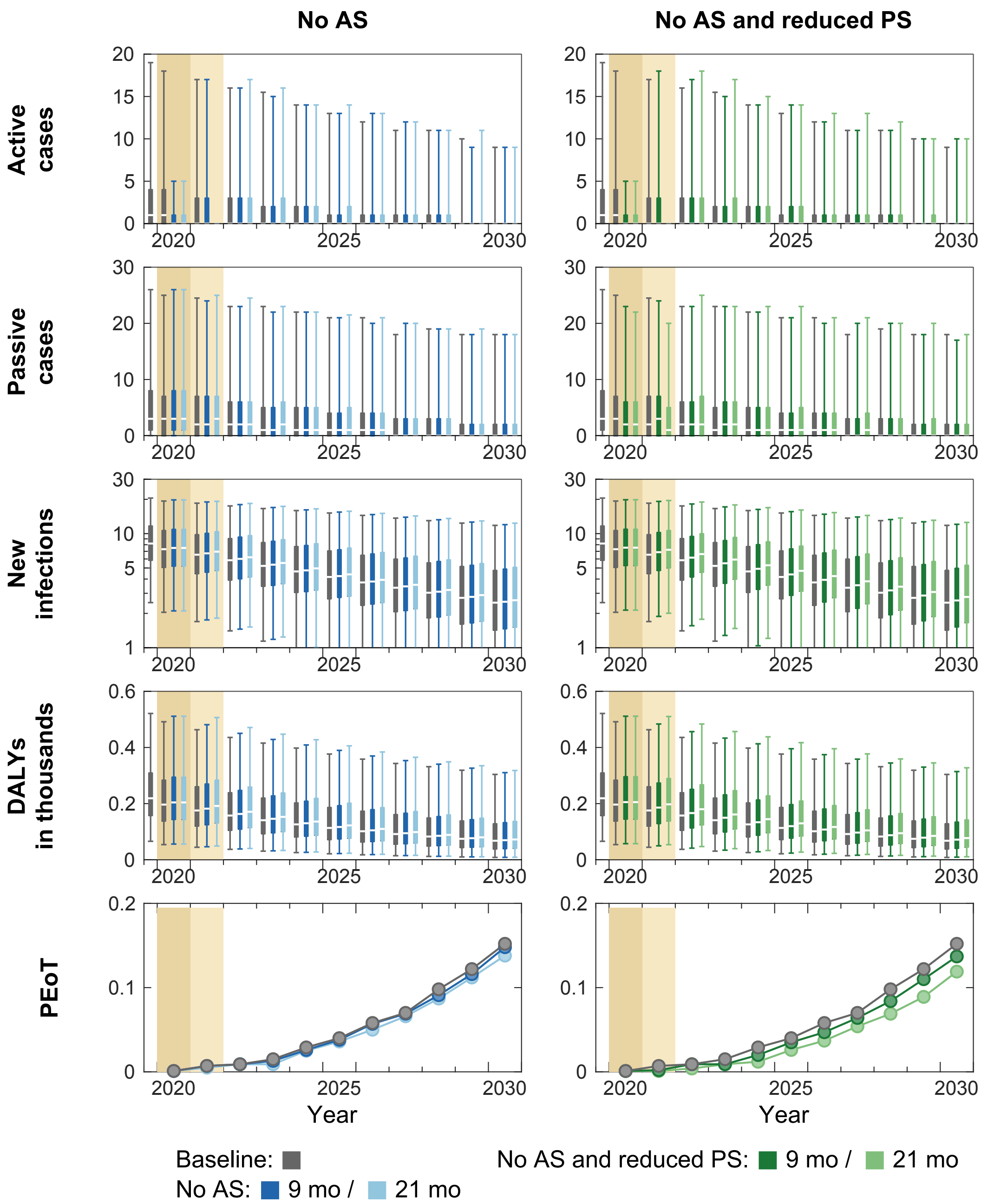


**Figure AA. Time series of model outputs in Lusanga health zone under the baseline and six interruption scenarios.** During 2014–2018, an average of 8% of the population participated in active screening resulting in 0.23 reported cases (from both active and passive screenings) per 10,000 per annum in Lusanga health zone. There are n = 10,000 independent samples, 10 from each of 1,000 independent samples from the joint posterior distributions of the fitted model parameters. Box plots summarise parameter and observational uncertainty. The lines in the boxes represent the medians of predicted results. The lower and upper bounds of the boxes indicate 25th and 75th percentiles. The minimum and maximum values are 2.5th and 97.5th percentiles and therefore whiskers cover 95% prediction intervals.
AS: active screening; PS: passive screening; VC: vector control; DALYs: disability-adjusted life years; PEoT: probability of elimination of transmission

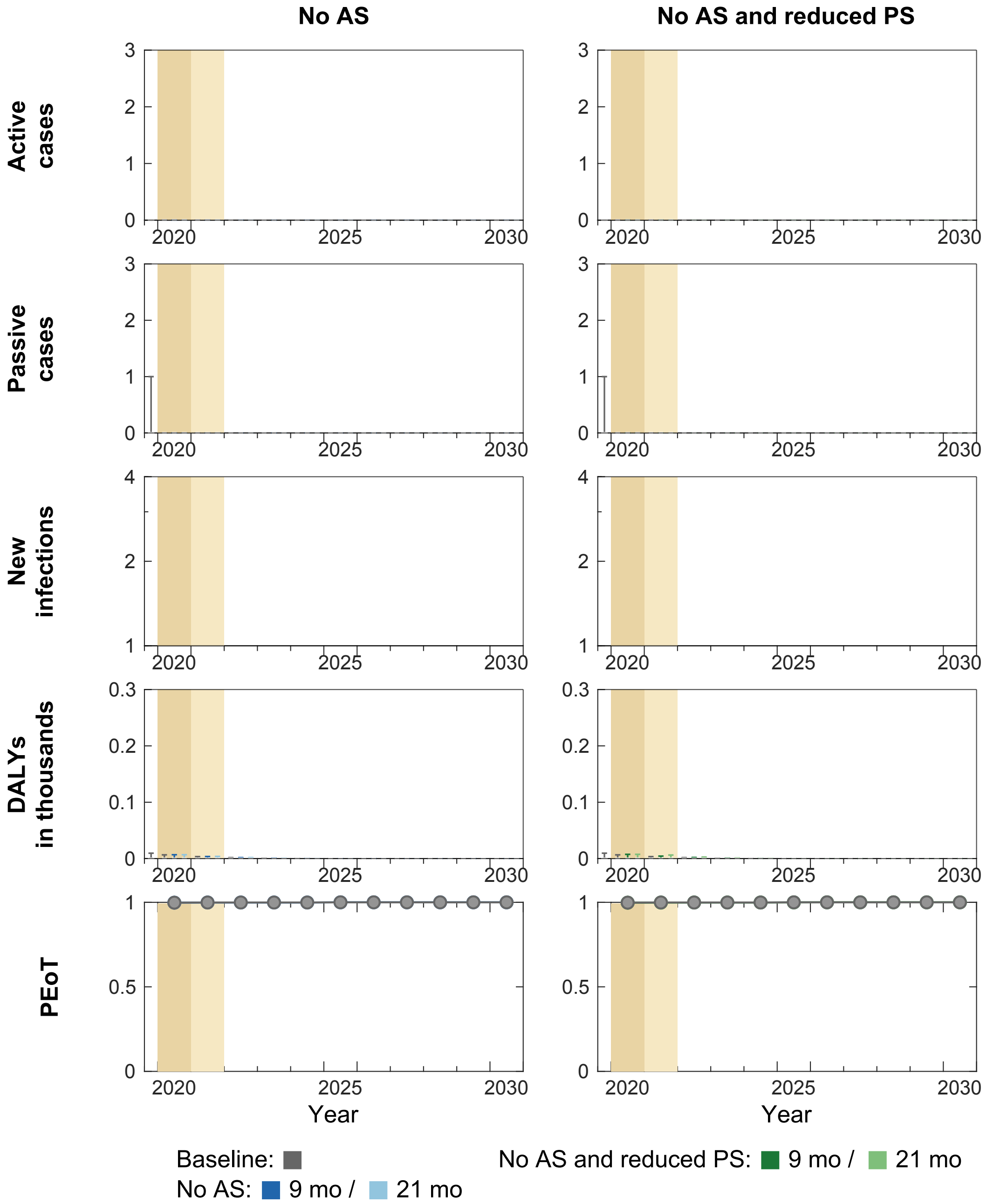


**Figure AB. Time series of model outputs in Moanza health zone under the baseline and six interruption scenarios.** During 2014–2018, an average of 8% of the population participated in active screening resulting in 0.21 reported cases (from both active and passive screenings) per 10,000 per annum in Moanza health zone. There are n = 10,000 independent samples, 10 from each of 1,000 independent samples from the joint posterior distributions of the fitted model parameters. Box plots summarise parameter and observational uncertainty. The lines in the boxes represent the medians of predicted results. The lower and upper bounds of the boxes indicate 25th and 75th percentiles. The minimum and maximum values are 2.5th and 97.5th percentiles and therefore whiskers cover 95% prediction intervals.
AS: active screening; PS: passive screening; VC: vector control; DALYs: disability-adjusted life years; PEoT: probability of elimination of transmission

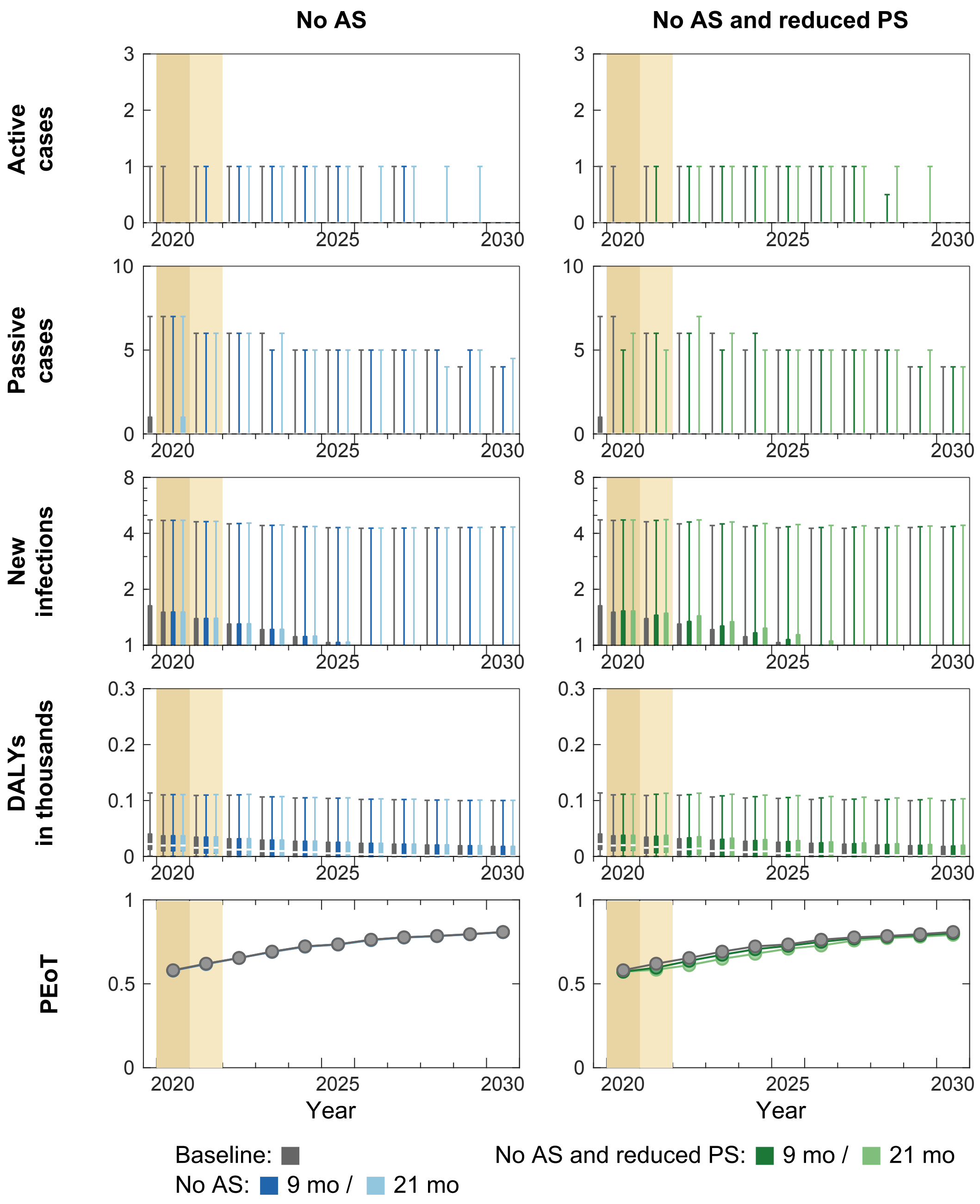


**Figure AC. Time series of model outputs in Mungindu health zone under the baseline and six interruption scenarios.** During 2014–2018, an average of 2% of the population participated in active screening resulting in 0.11 reported cases (from both active and passive screenings) per 10,000 per annum in Mungindu health zone. There are n = 10,000 independent samples, 10 from each of 1,000 independent samples from the joint posterior distributions of the fitted model parameters. Box plots summarise parameter and observational uncertainty. The lines in the boxes represent the medians of predicted results. The lower and upper bounds of the boxes indicate 25th and 75th percentiles. The minimum and maximum values are 2.5th and 97.5th percentiles and therefore whiskers cover 95% prediction intervals.

AS: active screening; PS: passive screening; VC: vector control; DALYs: disability-adjusted life years; PEoT: probability of elimination of transmission

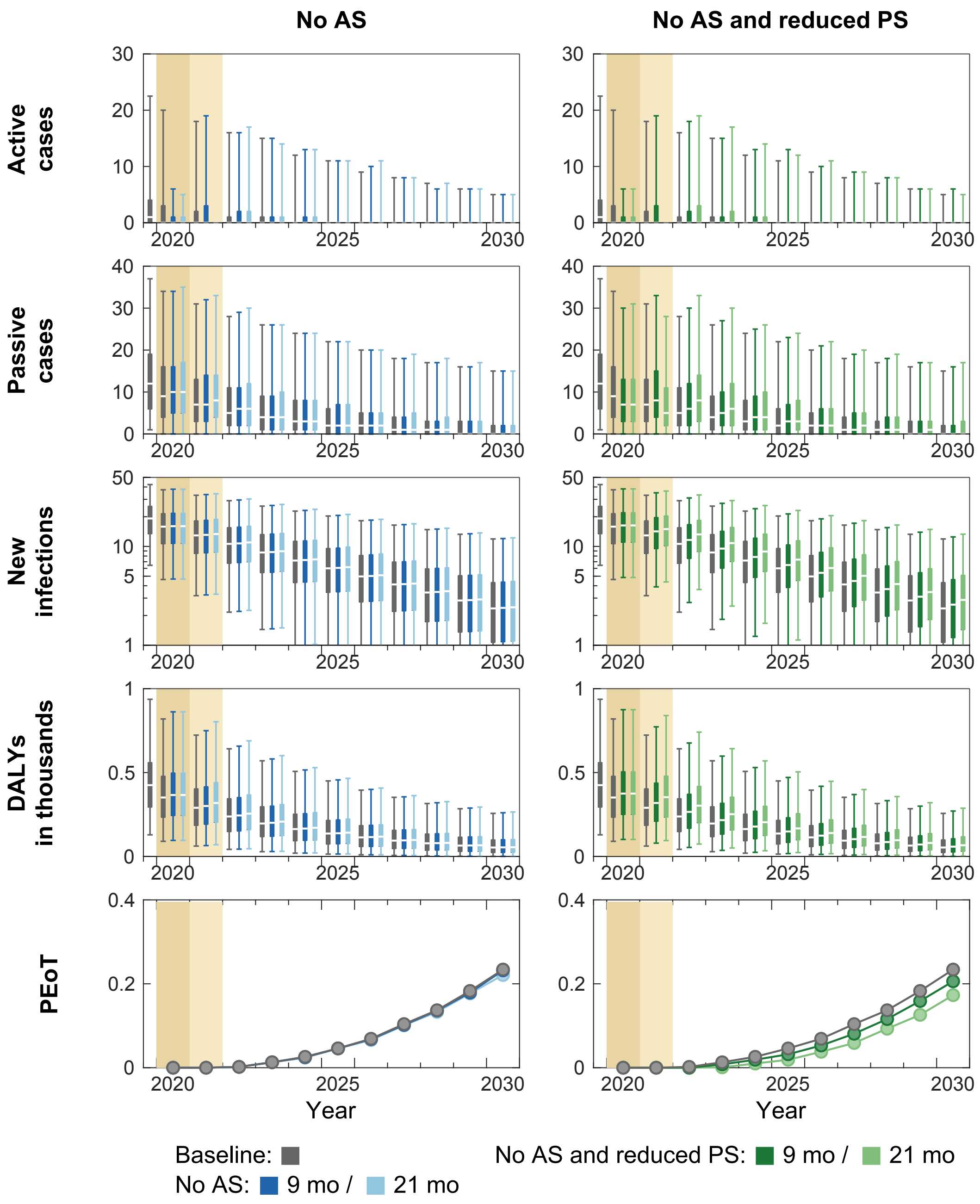


**Figure AD. Time series of model outputs in Nioki health zone under the baseline and six interruption scenarios.** During 2014–2018, an average of 16% of the population participated in active screening resulting in 2.82 reported cases (from both active and passive screenings) per 10,000 per annum in Nioki health zone. There are n = 10,000 independent samples, 10 from each of 1,000 independent samples from the joint posterior distributions of the fitted model parameters. Box plots summarise parameter and observational uncertainty. The lines in the boxes represent the medians of predicted results. The lower and upper bounds of the boxes indicate 25th and 75th percentiles. The minimum and maximum values are 2.5th and 97.5th percentiles and therefore whiskers cover 95% prediction intervals.
AS: active screening; PS: passive screening; VC: vector control; DALYs: disability-adjusted life years; PEoT: probability of elimination of transmission

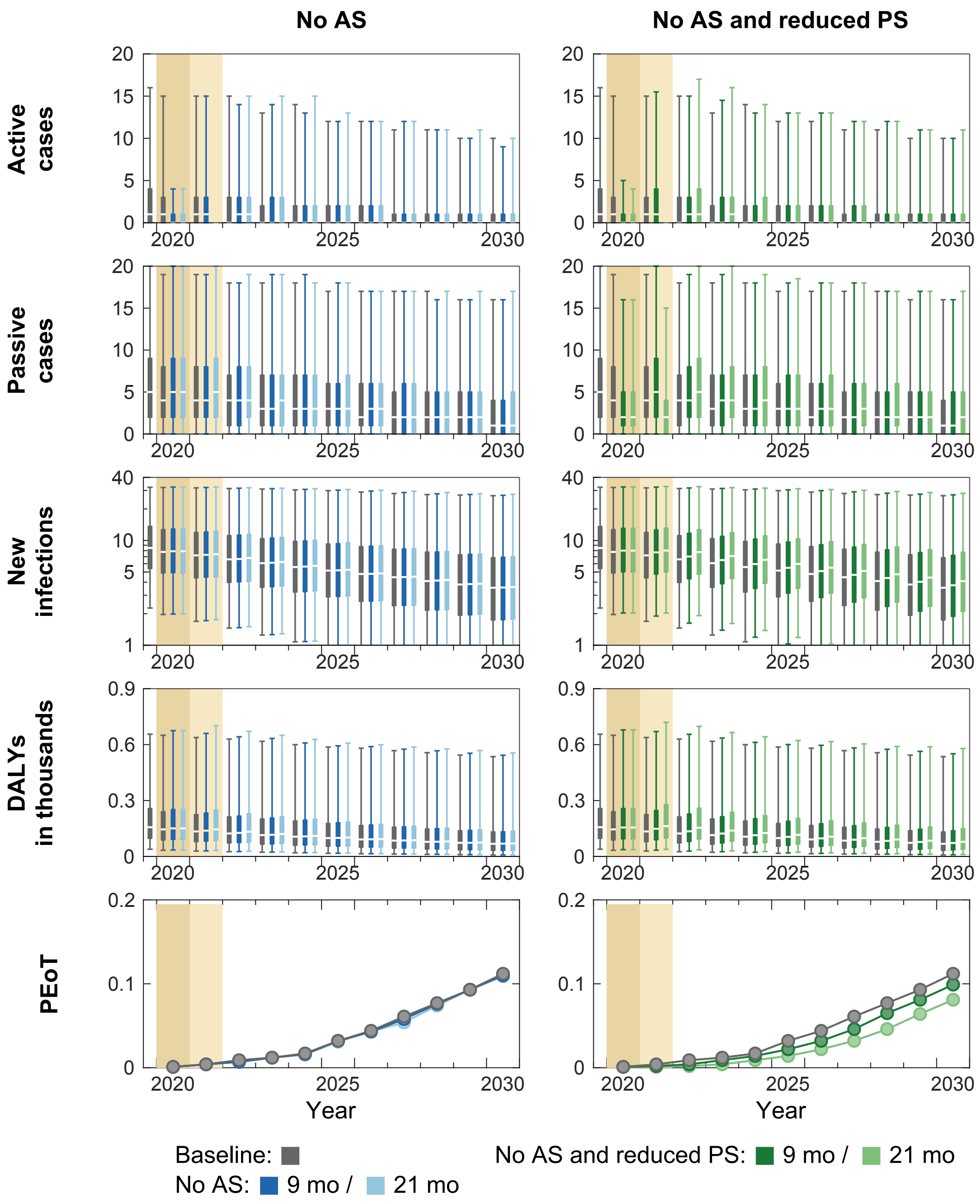


**Figure AE. Time series of model outputs in Ntand Embelo health zone under the baseline and six interruption scenarios.** During 2014–2018, an average of 16% of the population participated in active screening resulting in 2.69 reported cases (from both active and passive screenings) per 10,000 per annum in Ntand Embelo health zone. There are n = 10,000 independent samples, 10 from each of 1,000 independent samples from the joint posterior distributions of the fitted model parameters. Box plots summarise parameter and observational uncertainty. The lines in the boxes represent the medians of predicted results. The lower and upper bounds of the boxes indicate 25th and 75th percentiles. The minimum and maximum values are 2.5th and 97.5th percentiles and therefore whiskers cover 95% prediction intervals.

AS: active screening; PS: passive screening; VC: vector control; DALYs: disability-adjusted life years; PEoT: probability of elimination of transmission

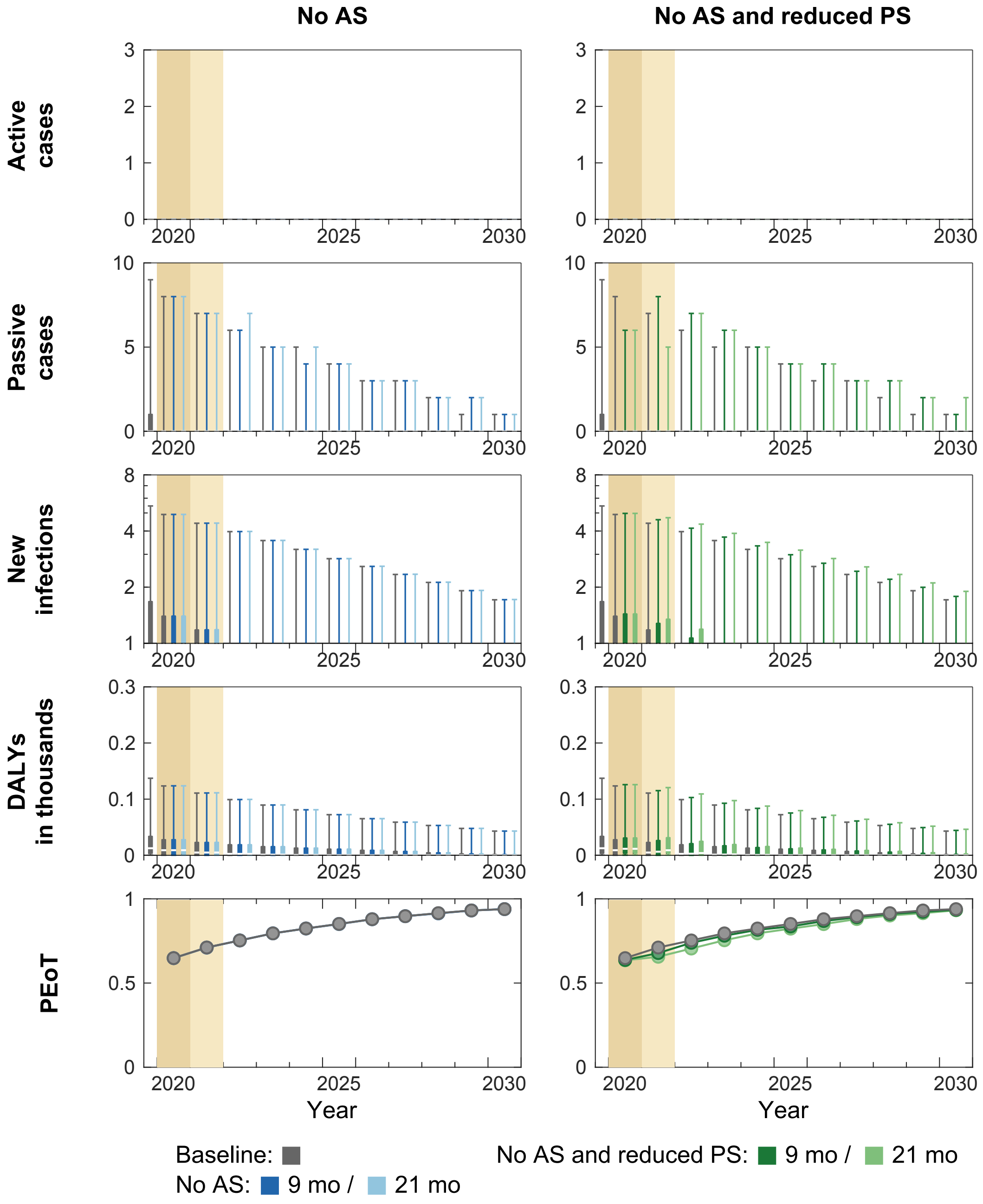


**Figure AF. Time series of model outputs in Oshwe health zone under the baseline and six interruption scenarios.** During 2014–2018, an average of 0% of the population participated in active screening resulting in 0.08 reported cases (from both active and passive screenings) per 10,000 per annum in Oshwe health zone. There are n = 10,000 independent samples, 10 from each of 1,000 independent samples from the joint posterior distributions of the fitted model parameters. Box plots summarise parameter and observational uncertainty. The lines in the boxes represent the medians of predicted results. The lower and upper bounds of the boxes indicate 25th and 75th percentiles. The minimum and maximum values are 2.5th and 97.5th percentiles and therefore whiskers cover 95% prediction intervals.
AS: active screening; PS: passive screening; VC: vector control; DALYs: disability-adjusted life years; PEoT: probability of elimination of transmission

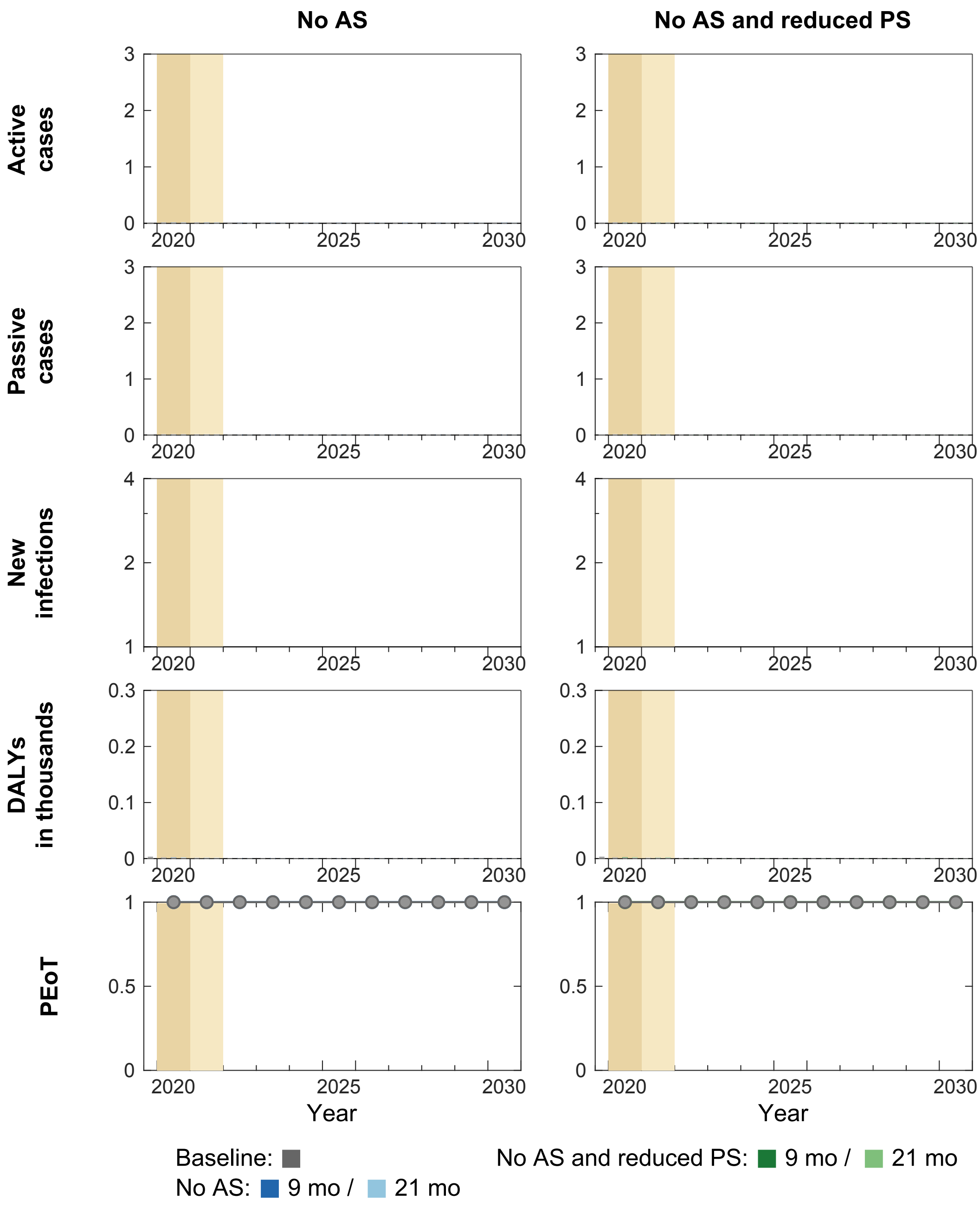


**Figure AG. Time series of model outputs in Pay Kongila health zone under the baseline and six interruption scenarios.** During 2014–2018, an average of 0% of the population participated in active screening resulting in 0.02 reported cases (from both active and passive screenings) per 10,000 per annum in Pay Kongila health zone. There are n = 10,000 independent samples, 10 from each of 1,000 independent samples from the joint posterior distributions of the fitted model parameters. Box plots summarise parameter and observational uncertainty. The lines in the boxes represent the medians of predicted results. The lower and upper bounds of the boxes indicate 25th and 75th percentiles. The minimum and maximum values are 2.5th and 97.5th percentiles and therefore whiskers cover 95% prediction intervals.
AS: active screening; PS: passive screening; VC: vector control; DALYs: disability-adjusted life years; PEoT: probability of elimination of transmission

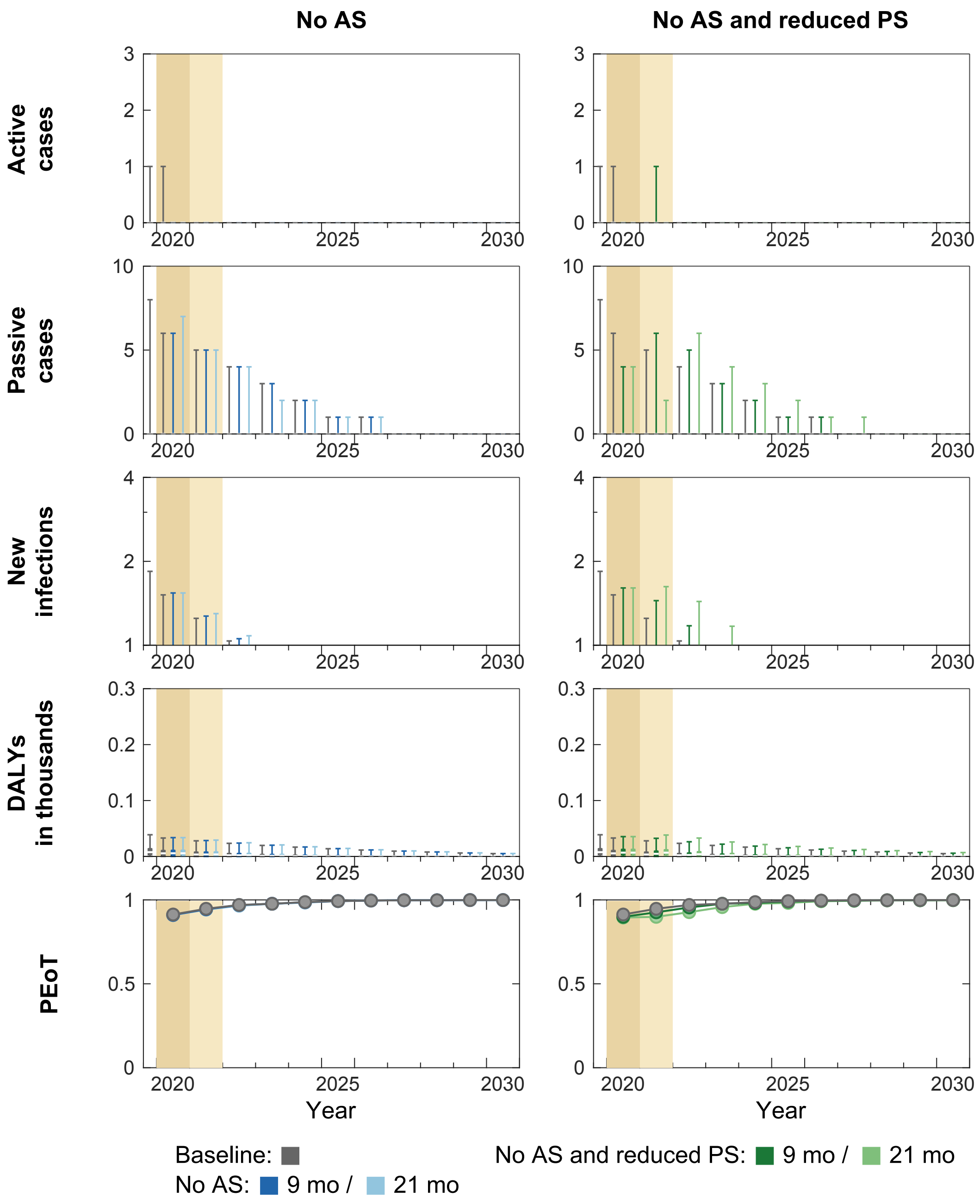


**Figure AH. Time series of model outputs in Popokabaka health zone under the baseline and six interruption scenarios.** During 2014–2018, an average of 4% of the population participated in active screening resulting in 0.53 reported cases (from both active and passive screenings) per 10,000 per annum in Popokabaka health zone. There are n = 10,000 independent samples, 10 from each of 1,000 independent samples from the joint posterior distributions of the fitted model parameters. Box plots summarise parameter and observational uncertainty. The lines in the boxes represent the medians of predicted results. The lower and upper bounds of the boxes indicate 25th and 75th percentiles. The minimum and maximum values are 2.5th and 97.5th percentiles and therefore whiskers cover 95% prediction intervals.
AS: active screening; PS: passive screening; VC: vector control; DALYs: disability-adjusted life years; PEoT: probability of elimination of transmission

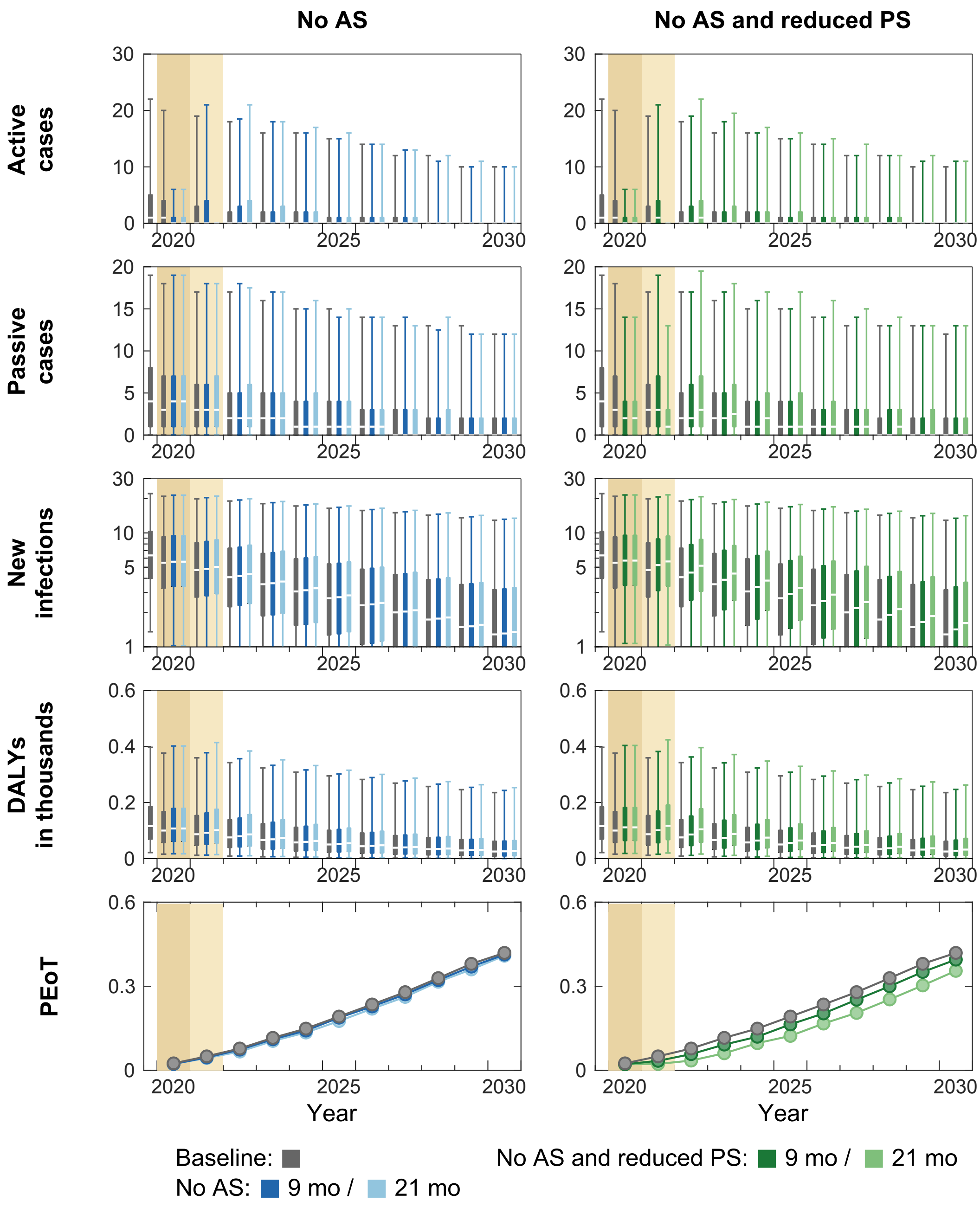


**Figure AI. Time series of model outputs in Sia health zone under the baseline and six interruption scenarios.** During 2014–2018, an average of 19% of the population participated in active screening resulting in 1.02 reported cases (from both active and passive screenings) per 10,000 per annum in Sia health zone. There are n = 10,000 independent samples, 10 from each of 1,000 independent samples from the joint posterior distributions of the fitted model parameters. Box plots summarise parameter and observational uncertainty. The lines in the boxes represent the medians of predicted results. The lower and upper bounds of the boxes indicate 25th and 75th percentiles. The minimum and maximum values are 2.5th and 97.5th percentiles and therefore whiskers cover 95% prediction intervals.
AS: active screening; PS: passive screening; VC: vector control; DALYs: disability-adjusted life years; PEoT: probability of elimination of transmission

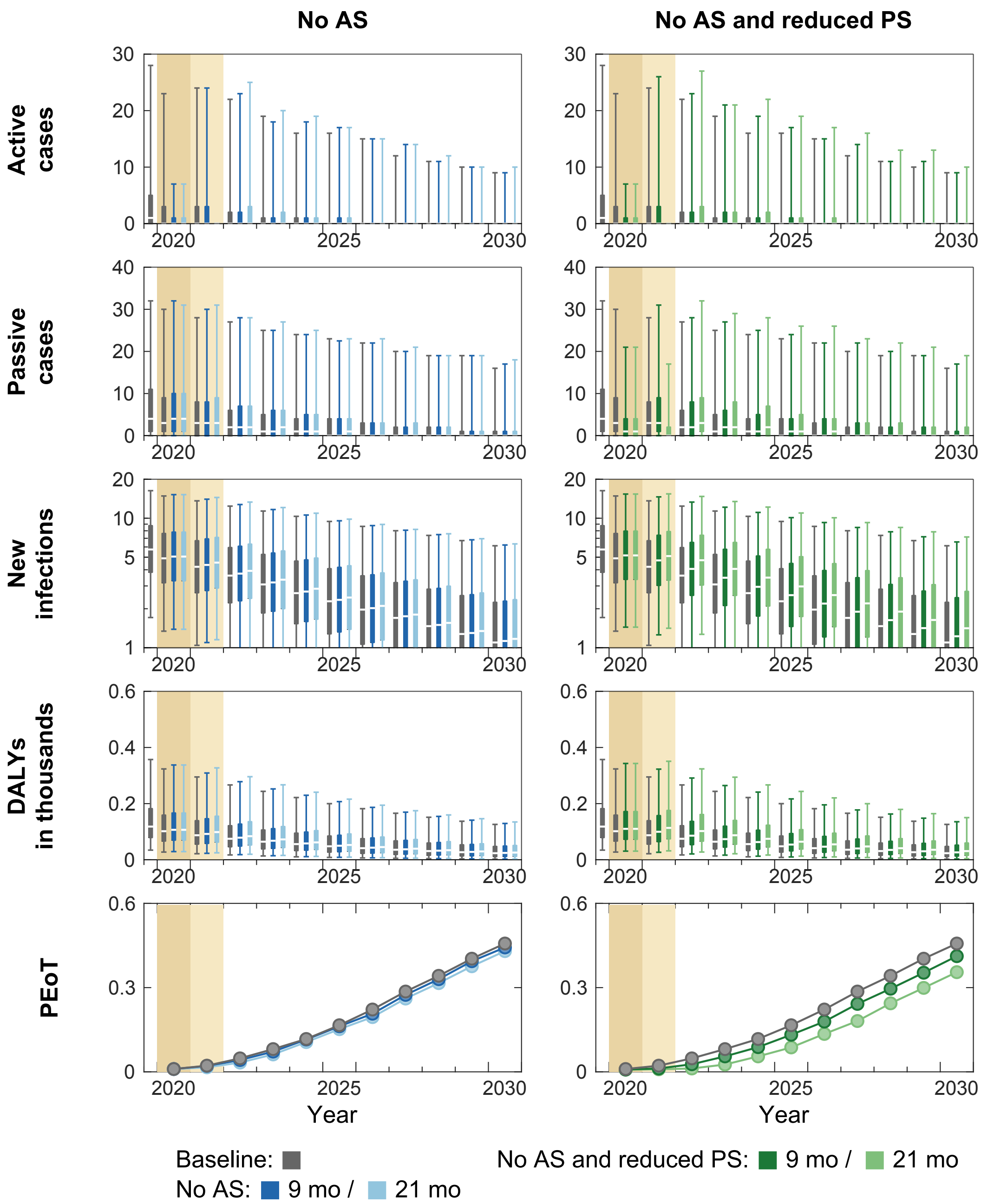


**Figure AJ. Time series of model outputs in Vanga health zone under the baseline and six interruption scenarios.** During 2014–2018, an average of 12% of the population participated in active screening resulting in 0.42 reported cases (from both active and passive screenings) per 10,000 per annum in Vanga health zone. There are n = 10,000 independent samples, 10 from each of 1,000 independent samples from the joint posterior distributions of the fitted model parameters. Box plots summarise parameter and observational uncertainty. The lines in the boxes represent the medians of predicted results. The lower and upper bounds of the boxes indicate 25th and 75th percentiles. The minimum and maximum values are 2.5th and 97.5th percentiles and therefore whiskers cover 95% prediction intervals.
AS: active screening; PS: passive screening; VC: vector control; DALYs: disability-adjusted life years; PEoT: probability of elimination of transmission

| Health zone | YEoT under baseline | Median delay in YEoT (in years) | | | | | |
|---|---|---|---|---|---|---|---|
| | | No AS | | No AS and reduced PS | | No AS or VC and reduced PS | |
| | | 9 months | 21 months | 9 months | 21 months | 9 months | 21 months |
| Yasa Bonga | 2017 | 0 | 0 | 0 | 0 | 0 | 0 |
| Masi Manimba | 2021 | 0 | 0 | 0 | 0 | 1 | 2 |
| Bandundu | 2020 | 0 | 0 | 0 | 0 | 1 | 2 |
| Kikongo | 2021 | 0 | 0 | 0 | 0 | 1 | 2 |
| Kwamouth | 2022 | 0 | 0 | 0 | 0 | 1 | 2 |
| Bokoro | 2023 | 0 | 0 | 0 | 0 | 0 | 1 |
| Bolobo | 2021 | 0 | 0 | 0 | 0 | 1 | 2 |
| Bulungu | 2022 | 0 | 0 | 0 | 0 | 1 | 2 |
| Mokala | 2023 | 0 | 0 | 0 | 0 | 0 | 1 |
| Mushie | 2023 | 0 | 0 | 0 | 0 | 0 | 1 |
| Yumbi | 2022 | 0 | 0 | 0 | 0 | 0 | 1 |
| Bagata | 2039 | 0 | 0 | 1 | 1 | | |
| Bandjau | >2050 | 0 | 0 | 1 | 2 | | |
| Boko | 2018 | 0 | 0 | 0 | 0 | | |
| Bosobe | 2022 | 0 | 0 | 0 | 1 | | |
| Djuma | 2036 | 0 | 0 | 1 | 2 | | |
| Idiofa | 2047 | 0 | 0 | 1 | 2 | | |
| Inongo | 2019 | 0 | 0 | 0 | 0 | | |
| Ipamu | 2038 | 0 | 0 | 1 | 2 | | |
| Kasongolunda | 2015 | 0 | 0 | 0 | 0 | | |
| Kenge | 2040 | 0 | 0 | 1 | 1 | | |
| Kikwit Nord | 2020 | 0 | 0 | 0 | 0 | | |
| Kikwit Sud | 2015 | 0 | 0 | 0 | 0 | | |
| Kimbau | 2011 | 0 | 0 | 0 | 0 | | |
| Kimputu | 2045 | 0 | 1 | 1 | 2 | Same as No AS and reduced PS | |
| Kiri | 2018 | 0 | 0 | 0 | 0 | | |
| Koshibanda | 2025 | 0 | 0 | 0 | 1 | | |
| Lusanga | 2040 | 0 | 1 | 1 | 1 | | |
| Moanza | 2013 | 0 | 0 | 0 | 0 | | |
| Mosango | 2027 | 0 | 1 | 1 | 1 | | |
| Mungindu | 2019 | 0 | 0 | 0 | 0 | | |
| Nioki | 2035 | 0 | 0 | 0 | 1 | | |
| Ntand Embelo | 2048 | 0 | 0 | 1 | 2 | | |
| Oshwe | 2019 | 0 | 0 | 0 | 0 | | |
| Pay Kongila | 2011 | 0 | 0 | 0 | 0 | | |
| Popokabaka | 2017 | 0 | 0 | 0 | 0 | | |
| Sia | 2032 | 0 | 0 | 1 | 2 | | |
| Vanga | 2031 | 0 | 0 | 1 | 2 | | |

**Table A. Expected year of elimination of transmission (YEoT) and delay to YEoT under interruption scenarios for all health zones of the former Bandundu province.** Blue shading represents health zones with on-going vector control (VC) before 2020, yellow shading represents those with planned VC, and orange represents those with no planned or on-going VC. In the health zones with no planned or on-going VC the *No AS or VC and reduced PS* scenario is identical to *No AS and reduced PS*.
AS: active screening; PS: passive screening; VC: vector control; YEoT: year of elimination of transmission

| Health zone | Median DALYs under baseline | Additional DALYs accrued (median) | | | | | |
|---|---|---|---|---|---|---|---|
| | | No AS | | No AS and reduced PS | | No AS or VC and reduced PS | |
| | | 9 months | 21 months | 9 months | 21 months | 9 months | 21 months |
| Yasa Bonga | 47.0 | <0.1 | 0.1 | 10.9 | 22.3 | 10.9 | 22.3 |
| Masi Manimba | 868.8 | 56.1 | 103.3 | 144.4 | 289.5 | 165.4 | 369.6 |
| Bandundu | 182.7 | 17.3 | 27.8 | 46.6 | 90.7 | 56.1 | 123.4 |
| Kikongo | 1131.1 | 17.7 | 43.9 | 84.6 | 182.6 | 92.1 | 218.1 |
| Kwamouth | 1445.9 | 160.4 | 268.8 | 365.9 | 727.5 | 463.7 | 1069.3 |
| Bokoro | 3221.4 | 22.7 | 58.8 | 190.5 | 476.5 | 190.5 | 631.4 |
| Bolobo | 148.7 | 15.0 | 26.2 | 64.7 | 129.2 | 106 | 232.2 |
| Bulungu | 1091.4 | 62.0 | 133.6 | 191.2 | 394.5 | 401.9 | 847.7 |
| Mokala | 1085.6 | 47.2 | 104.6 | 131.3 | 284.5 | 131.3 | 516.6 |
| Mushie | 1516.2 | 50.4 | 112.6 | 117.0 | 249.8 | 117.0 | 538.7 |
| Yumbi | 495.8 | 14.0 | 32.3 | 52.6 | 120.3 | 52.6 | 166.2 |
| Bagata | 2421.3 | 92.0 | 216.4 | 303.5 | 652.8 | | |
| Bandjau | 3433.8 | 16.0 | 33.0 | 32.3 | 76.5 | | |
| Boko | 18.5 | 0.3 | 0.5 | 3.1 | 6.5 | | |
| Bosobe | 167.5 | 9.4 | 18.9 | 19.0 | 43.9 | | |
| Djuma | 1371.1 | 74.4 | 176.3 | 199.9 | 451.2 | | |
| Idiofa | 1119.5 | 29.1 | 63.4 | 71.2 | 168.9 | | |
| Inongo | 27.6 | <0.1 | <0.1 | 8.8 | 24.8 | | |
| Ipamu | 1151.5 | 39.2 | 76.3 | 141.9 | 332.9 | | |
| Kasongolunda | 6.5 | <0.1 | <0.1 | 2.7 | 7.0 | | |
| Kenge | 1945.2 | 99.1 | 204.8 | 187.3 | 417.3 | | |
| Kikwit Nord | 22.3 | <0.1 | <0.1 | 10.5 | 27.9 | | |
| Kikwit Sud | 0.9 | <0.1 | <0.1 | 0.6 | 1.4 | | |
| Kimbau | <0.1 | <0.1 | <0.1 | <0.1 | <0.1 | Same as No AS and reduced PS | |
| Kimputu | 3376.2 | 197.4 | 404.6 | 357.0 | 774.2 | | |
| Kiri | 42.2 | <0.1 | <0.1 | 3.9 | 7.6 | | |
| Koshibanda | 255.0 | 5.1 | 13.0 | 15.3 | 38.8 | | |
| Lusanga | 1326.7 | 44.4 | 97.4 | 78.0 | 172.1 | | |
| Moanza | <0.1 | <0.1 | <0.1 | <0.1 | <0.1 | | |
| Mosango | 322.8 | 35.2 | 76.3 | 61.4 | 152.7 | | |
| Mungindu | 81.3 | 0.3 | 0.6 | 9.3 | 17.2 | | |
| Nioki | 1769.6 | 47.3 | 96.7 | 169.4 | 401.3 | | |
| Ntand Embelo | 1125.9 | 14.9 | 34.0 | 72.1 | 180.8 | | |
| Oshwe | 18.7 | <0.1 | <0.1 | 4.3 | 8.4 | | |
| Pay Kongila | <0.1 | <0.1 | <0.1 | <0.1 | <0.1 | | |
| Popokabaka | 14.1 | 0.3 | 0.5 | 3.6 | 8.0 | | |
| Sia | 610.3 | 22.0 | 55.7 | 67.7 | 163.0 | | |
| Vanga | 593.2 | 28.1 | 61.8 | 75.4 | 172.0 | | |

**Table B. Total median disability-adjusted life years (DALYs) under baseline and additional DALYs accrued under interruption scenarios between 2020 and 2030 for all health zones of the former Bandundu province.** Blue shading represents health zones with on-going vector control (VC) before 2020, yellow shading represents those with planned VC, and orange represents those with no planned or on-going VC. In the health zones with no planned or on-going VC the *No AS or VC, and reduced PS* scenario is identical to *No AS and reduced PS*.

AS: active screening; PS: passive screening; VC: vector control; DALYs: disability-adjusted life years

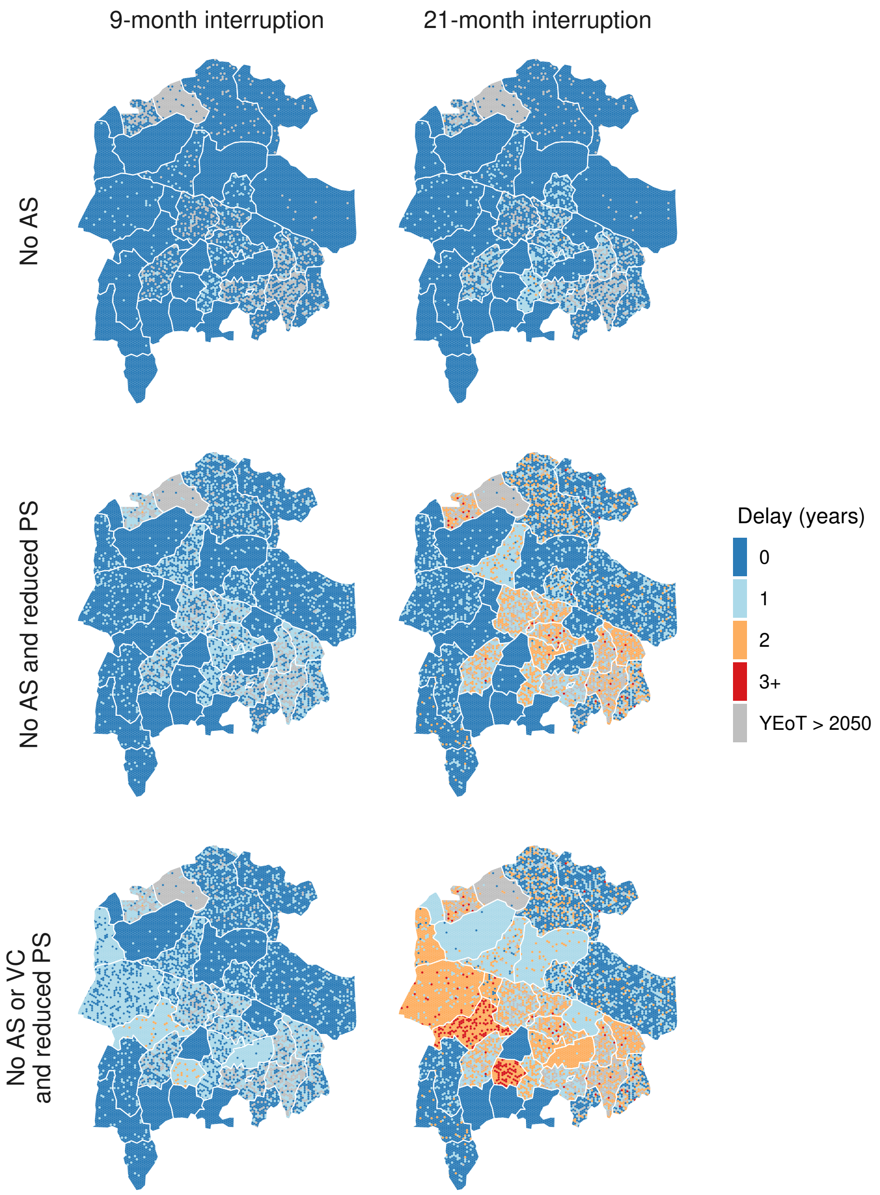


**Figure AK. Representation of within-health zone distribution of delay in YEoT under six interruption scenarios.** Colours of individual hexagons represent a random sample from the posterior distribution for that health zone. 'YEoT > 2050' where the realised YEoT, either baseline, interruption, or both, was beyond 2050. In each health zone, the baseline strategy is either *MeanAS* or *MeanAS+VC* depending on its VC status (summarised in Table 1 in the main text). Shapefiles used to produce these maps were provided by Nicole Hoff and Cyrus Sinai under a CC-BY licence (current versions can be found at https://data.humdata.org/dataset/drc-health-data).
AS: active screening; PS: passive screening; VC: vector control; YEoT: year of elimination of transmission
