## Supplementary material for "A modelling assessment of short- and medium-term risks of programme interruptions for *gambiense* human African trypanosomiasis in the DRC": S3 Text: PRIME-NTD criteria

### Supporting information – S3 Text: PRIME-NTD criteria

**Ching-I Huang[1,2,+,*], Ronald E Crump[1,2,3], Emily H Crowley[1,2], Andrew Hope[4], Paul R Bessell[5], Chansy Shampa[6], Erick Mwamba Miaka[6], and Kat S Rock[1,2,+]**

[1]Zeeman Institute for System Biology and Infectious Disease Epidemiology Research, The University of Warwick, Coventry, U.K
[2]Mathematics Institute, The University of Warwick, Coventry, U.K.
[3]The School of Life Sciences, The University of Warwick, Coventry, U.K.
[4]Liverpool School of Tropical Medicine (LSTM), Liverpool, U.K.
[5]Independent Consultant, Edinburgh, U.K.
[6]Programme National de Lutte contre la Trypanosomiase Humaine Africaine (PNLTHA), Kinshasa, D.R.C.
[*]
[+]these authors contributed equally to this work

#### PRIME-NTD criteria

It has been recommended that good modelling practises should meet the five key principles relating to communication, quality and relevance of analyses - known as Policy-Relevant Items for Reporting Models in Epidemiology of Neglected Tropical Diseases (PRIME-NTD)[1]. We demonstrate how these PRIME-NTD criteria have each been addressed in Table A.

| | What has been done to satisfy the principle? | Where in the manuscript is this described? |
|---|---|---|
| 1. Stakeholder engagement | This modelling study has been conducted in conjunction with a range of partners, including the national sleeping sickness programme of DRC (PNLTHA-DRC) – co-authors E M Miaka and C Shampa, as well as experts in tsetse control – co-author A Hope. Numerous discussions were had to ensure the modellers produced meaningful outputs with policy relevance. | Authorship list |
| 2. Complete model documentation | Full model fitting code and documentation is available through OpenScienceFramework (OSF). The model is fully described in the main text and supplementary information. Previous versions of the model are fully described elsewhere[2,3]. | See Methods section in the main text, S1 Methods and at OSF (https://osf.io/gur7c/) |
| 3. Complete description of data used | The original data and how we aggregated the data for fitting were described in a previous model fitting paper[2] and can be viewed through the corresponding graphical user interface https://hatmepp.warwick.ac.uk/fitting/v2/. In the present study we don't update these fits, but use the posterior parameters to generate new future projections. | See Materials and Methods section and previous publication[2] |
| 4. Communicating uncertainty | *Parameter uncertainty*: We used estimates from joint posterior distributions of fitted parameters generated from the previous model fitting.<br>*Prediction uncertainty*: We represent uncertainty in our results by providing box and whisker plots for predicted outcomes (median, 50% and 95% credible intervals) or probabilities of outcomes. | *Parameter uncertainty*: All results figures and tables<br><br>*Prediction uncertainty*: Figs 2–4, and S2 Text Figs A–AK |
| 5. Testable model outcomes | For this analysis we used AS coverage for 2000–2018 but at the time of the analysis 2019 onward was unavailable which could have impacted our results. We had assumed this missing coverage data for 2019 was the mean of 2014–2018. When the data becomes available for a few years after 2020 (e.g. until the end of 2022), it would be possible to perform modelling validation to examine our predictions by using the actual screening numbers with the same model fits and compare to the case reporting observed during and post interruption. Our results presented in this article and our open source code mean this validation exercise should be straightforward to conduct. It would also be possible to refit the model to the actual screening and case data to quantify if there is any statistical evidence of the interruption's impact on PS during 2020. | See main text results |

**Table A.** PRIME-NTD criteria fulfillment. We summarise how the NTD Modelling Consortium's "5 key principles of good modelling practice" have been met in the present study.
